# MITIGATING THE 4^th^ WAVE OF THE COVID-19 PANDEMIC IN ONTARIO

**DOI:** 10.1101/2021.09.02.21263000

**Authors:** Lauren E. Cipriano, Wael M. R. Haddara, Beate Sander

## Abstract

**Background:** The goal of this study was to project the number of COVID-19 cases and demand for acute hospital resources for Fall of 2021 in a representative mid-sized community in southwestern Ontario. We sought to evaluate whether current levels of vaccine coverage and contact reduction could mitigate a potential 4^th^ wave fueled by the Delta variant, or whether the reinstitution of more intense public health measures will be required.

**Methods:** We developed an age-stratified dynamic transmission model of COVID-19 in a mid-sized city (population 500,000) currently experiencing a relatively low, but increasing, infection rate in Step 3 of Ontario’s Wave 3 recovery. We parameterized the model using the medical literature, grey literature, and government reports. We estimated the current level of contact reduction by model calibration to cases and hospitalizations. We projected the number of infections, number of hospitalizations, and the time to re-instate high intensity public health measures over the fall of 2021 under different levels of vaccine coverage and contact reduction.

**Results:** Maintaining contact reductions at the current level, estimated to be a 17% reduction compared to pre-pandemic contact levels, results in COVID-related admissions exceeding 20% of pre-pandemic critical care capacity by late October, leading to cancellation of elective surgeries and other non-COVID health services. At high levels of vaccination and relatively high levels of mask wearing, a moderate additional effort to reduce contacts (30% reduction compared to pre-pandemic contact levels), is necessary to avoid re-instating intensive public health measures. Compared to prior waves, the age distribution of both cases and hospitalizations shifts younger and the estimated number of pediatric critical care hospitalizations may substantially exceed 20% of capacity.

**Discussion:** High rates of vaccination coverage in people over the age of 12 and mask wearing in public settings will not be sufficient to prevent an overwhelming resurgence of COVID-19 in the Fall of 2021. Our analysis indicates that immediate moderate public health measures can prevent the necessity for more intense and disruptive measures later.

## INTRODUCTION

Each successive wave of the COVID-19 pandemic raises different public health challenges. In Ontario, Canada, a significant 3^rd^ wave in the spring of 2021, driven by the Alpha variant, led to an extended stay-at-home order and closure of schools [1]. Concurrent with a large vaccination campaign, gradual release of restrictions has kept the incidence rate low through the summer. However, the more infectious and more severe Delta variant has become the dominant strain circulating across Ontario and cases are increasing [2, 3].

In other regions, including those with similarly high rates of vaccination, the Delta variant has led to rapidly increasing cases and significant pressure on health system resources [4-8]. Unique to this current wave is the lower average age of hospitalized patients [8-10], explained by a combination of factors including increased transmissibility and disease severity of the Delta variant [7, 11-16] paired with generally high vaccine coverage rates in older adults and hence protection from severe disease [14, 17-23], lower vaccine coverage in younger adults, and the ineligibility of children under 12 years for vaccination [2, 3].

The goal of this study was to project the number of COVID-19 cases and demand for acute hospital resources for Fall of 2021 in a mid-sized community in southwestern Ontario (City of London and Middlesex County), with an approximate population of 500,000. We sought to evaluate whether current levels of vaccine coverage and contact reduction could mitigate a potential 4^th^ wave, or whether reinstitution of more intense public health measures is required. Specifically, we evaluated public health interventions in the context of the resumption of in-person K-12 and post-secondary education.

## METHODS

We extended a previously developed deterministic dynamic compartmental model of SARS-CoV-2 transmission that predicts health outcomes and resource use in a representative mid-sized city with a full-time population of 500,000 and an additional part-time academic year population of 50,000 post-secondary students [24]. We estimated model parameters, including the duration of time spent in each health state, the infectiousness of COVID-19, the probability of needing general and specialized hospital resources, COVID-19-associated mortality, the effectiveness of COVID-19 public health measures, and the effectiveness of vaccination using the peer-reviewed literature, pre-published reports, government reports, and expert opinion. Details about the model structure, implementation, and parameterization are provided in the **Supplemental Material**.

Scenarios presented throughout the analysis vary in the level of contact reduction compared to the pre-pandemic number of close contacts. We performed sensitivity analysis to explore the impact of parameter uncertainty on model projections.

### Model structure

The modelled population is divided into compartments based on age (0-4 years, 5-11 years, 12-17 years, 18-24 years, 25-49 years, 50-59 years, 60-69 years, and 70+ years) or by the primary setting of contacts (Post-secondary students and Long-term care (LTC) residents). A schematic of the COVID-19 health states is presented in **FIGURE 1**. In the model, susceptible individuals (vaccinated and unvaccinated) may become infected through interaction with infected individuals who may or may not be aware of their infection status. Infection has a pre-symptomatic phase in which an infected individual can transmit the infection to others [25-27]. Individuals may become aware of their infection status through symptom-based surveillance or contact tracing. Individuals aware of their infection status with mild or moderate symptoms may isolate at home reducing their contacts by 90%. Some patients develop severe symptoms requiring hospitalization or critical illness requiring admission to an intensive care unit (ICU). In the model, only hospitalized patients and LTC residents die of COVID-19 infection.

**Figure 1.**
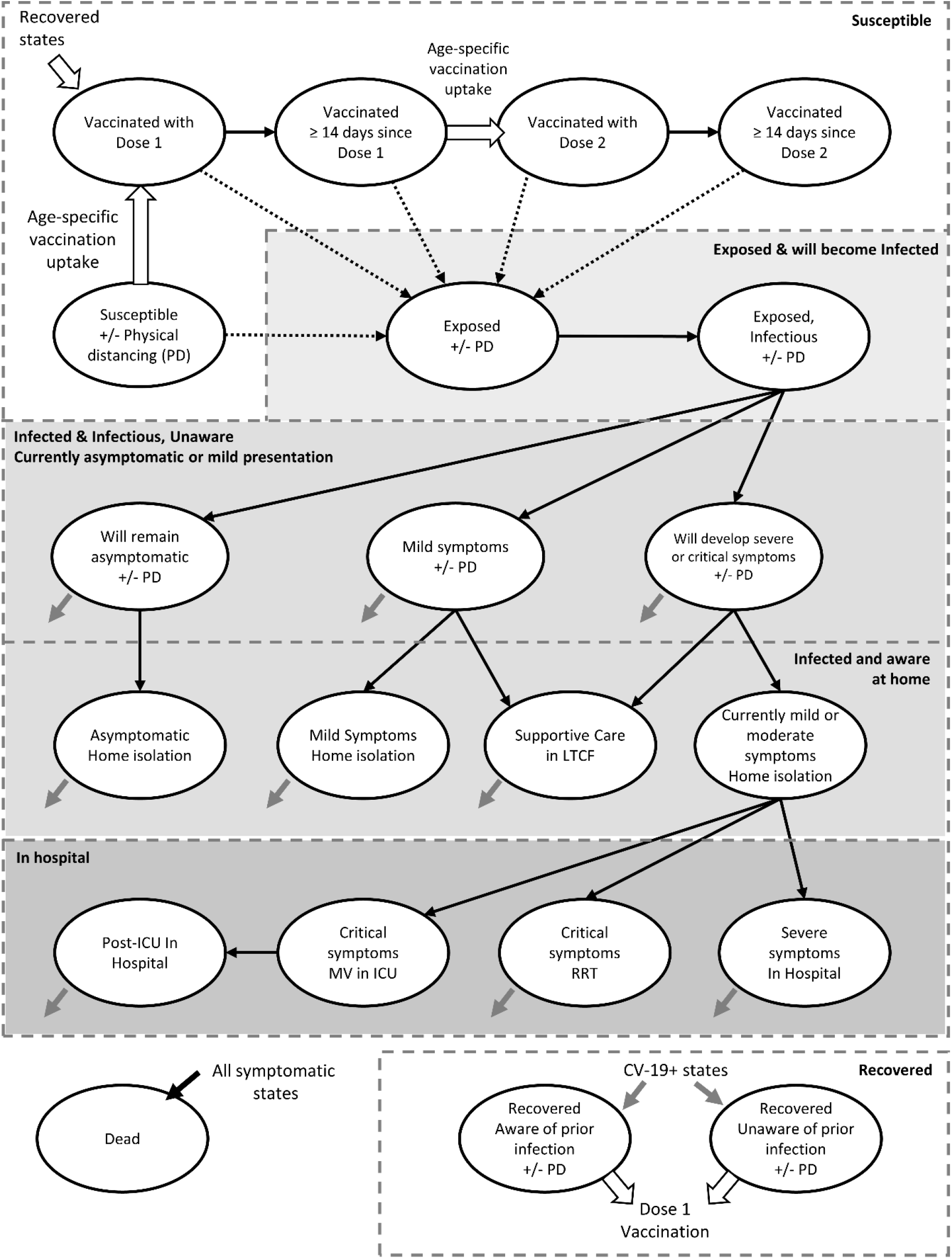
Model schematic.

### Impact of Variants of Concern

We estimated the prevalence of the Alpha (B.1.1.7) and Delta (B.1.617.2) variants in the community using Ontario public health surveillance reports [2] (**APPENDIX FIGURE 1**). We assumed the Alpha variant to be 1.5 times more infectious than the original strain dominant in North America in the spring of 2020 [16, 28, 29] and the Delta variant to be 1.5 times more infectious than Alpha [15, 16]. In addition, we assumed the overall probability of hospitalization is 1.63 times higher for infection with the Alpha variant [13, 30, 31], compared to the original strain, and an additional 1.85 times higher for infection with the Delta variant compared to Alpha [11-13].

### Contact mixing patterns

Age-specific contact mixing patterns were estimated using the POLYMOD study, adjusted to reflect the network structure of the Canadian population [32]. Additionally, we estimated children’s incremental school-based contacts using studies that used wireless sensor technologies to measure close contacts in schools [33, 34]. We included two special populations in the model, post-secondary students, whose contacts were estimated using a survey of university students [35], and LTC residents, whose contacts were estimated using wireless sensor technology in a Canadian long-term care facility [36, 37].

### Dynamic behaviour change in response to COVID-19 activity in the community

In some scenarios, we consider the impact of dynamic behaviour in response to COVID-19 outcomes in the community. These changes in behaviour are intended to capture individual decision making in the absence of or in advance of government intervention (e.g., reducing social contacts), such as those observed in a U.S. study [38], as well as policy changes that may be instituted in the community (e.g., capacity limits on retail services, stay-at-home orders). In the base case, we assumed that the community begins increasing the level of contact reduction by 0.75% per day once 7 ICU beds are occupied (10% of pre-pandemic capacity), and by an additional 1.5% per day after there have been 5 COVID-19 deaths in the past 10 days. The maximum level of contact reduction was set to 50% in adults and 70% in children and teenagers, representing, on the higher end of this range, the closure of schools.

### Masks

We assume children under 12 years of age, LTC residents, and ‘high-intensity physical distancers’ (75% of the population) wear masks for 86% of their contacts and that the remainder (‘low-intensity physical distancers’) wear masks for 38% of their contacts consistent with reported levels of mask wearing in a survey of Canadians [39]. We assume that as people become fully vaccinated, they reduce their mask wearing to 38% of their contacts. We assume cloth masks reduce the probability of disease transmission by 46% based on the reduction in transmission observed in German cities after the introduction of a mask mandate [40].

### Vaccination uptake and effectiveness

We implemented first and second dose uptake using actual age-specific weekly vaccination data published by the local health unit until the week of August 15, 2021 and estimate forward using a decreasing weekly rate of uptake (**APPENDIX FIGURE 2**).

In the base case, we assumed that full vaccination reduces the probability of infection by the Alpha variant by 90% [18, 41-45] and by the Delta variant by 76.5% (15% less than Alpha) [18, 23, 46]. To avoid double counting the benefit already conferred from the reduced risk of infection, we calculate the risk reduction for symptomatic disease and the risk reduction of hospitalization conditional on infection. This led us to assume a 45% reduction in the probability of symptomatic infection and a 60% reduction in the risk of hospitalization conditional on infection in vaccinated individuals. Cumulatively, through both the prevention of infections and reduced disease severity, in the model, vaccination reduces hospitalizations by 96% against the Alpha variant and 91% against the Delta variant, consistent with population level estimates of vaccine effectiveness [18-23, 44, 47].

## RESULTS

### Base case projections

Ontario entered Step 3 of the Wave 3 recovery plan on July 16, 2021 [48, 49]. We estimate the average level of contact reduction in Step 3 to be 17%, compared to pre-pandemic numbers of daily close contacts, via model calibration to diagnosed cases, hospitalizations, and deaths to August 25, 2021 (**APPENDIX FIGURE 3**). From August 25, 2021, onwards, we predict the number of new infections per day, ward bed occupancy, and ICU bed occupancy under different levels of contact reduction. At all levels of contact reduction, the number of new infections per day increases upon commencement of in-person schools and post-secondary education. When the level of contact reduction is between 15% and 20%, similar to the level of contact reduction estimated for August 2021, the number of infections per day exceeds 40 per 100,000 in late September. In contrast, at levels of contact reduction between 25% and 27.5%, the number of infections per day exceeds 40 per 100,000 nearly a month later, in mid-October to early-November. Correspondingly, ICU occupancy exceeds 15 COVID-19 patients (representing 20% of pre-pandemic capacity), the level at which elective surgeries and other procedures must be cancelled, in late October, mid-November, or early December for scenarios with contact reductions of 20%, 25%, and 27% relative to pre-pandemic average daily close contacts (**FIGURE 2**).

**Figure 2.**
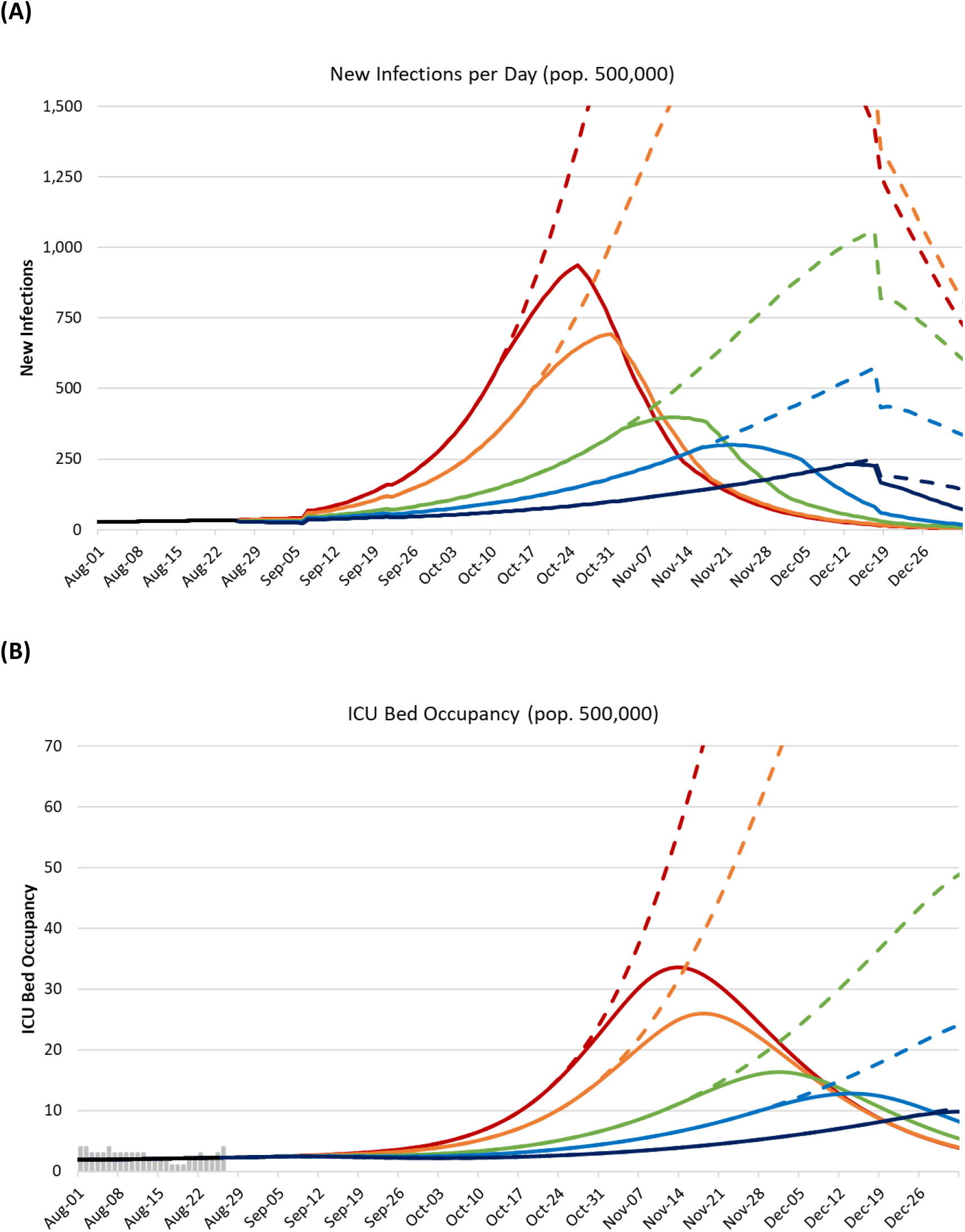
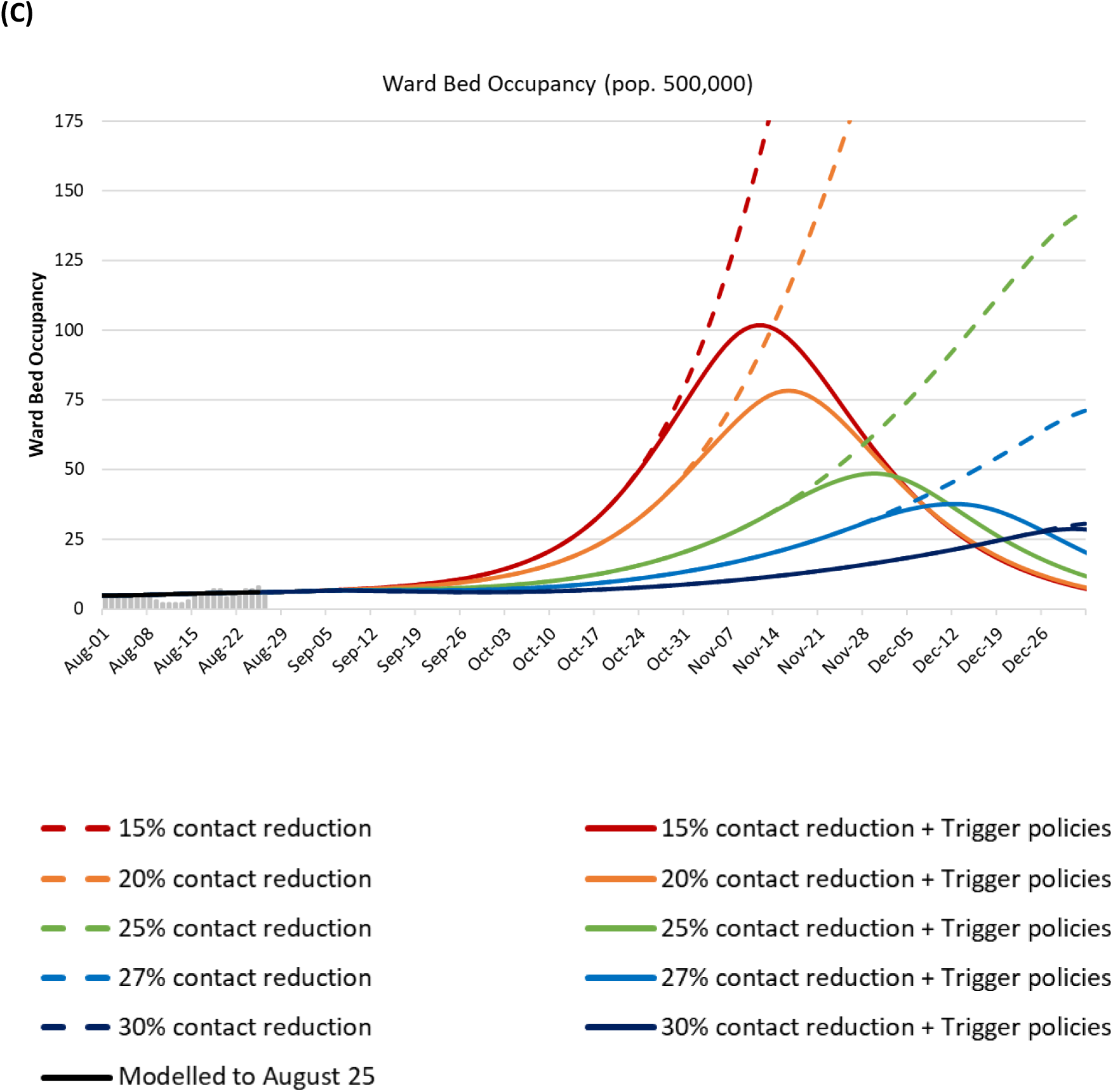
Projected (A) new infections per day, (B) ICU occupancy, and (C) ward bed occupancy for August 2021 through December 2021 in London-Middlesex in and from members of the local community without (dashed lines) and with (solid lines) incorporating individual-level behaviour change and the re-instatement of restrictive physical distancing policies and business closures. In these scenarios, behaviours and policies begin to change when the ICU occupancy reaches 7 people (10% of pre-COVID capacity) or there have been at least 5 COVID-19 deaths in 10 days.

We assumed that individuals change their contact behaviours and policy-makers will reinstitute restrictive public health measures to reduce contacts in response to worsening COVID-19 outcomes in the community. In the base case analysis, we assumed that contact reduction would begin to increase after 7 ICU beds were occupied with COVID-19 patients (representing 10% of pre-COVID capacity) and, more intensely, when there were 5 COVID-19 deaths within 10 days in the community. In scenarios with 15%, 20%, and 25% contact reduction, the ICU threshold, of 7 beds, was exceeded October 12^th^, October 18^th^, and November 3^rd^ respectively, and the number of COVID-19 deaths exceeded the threshold of 5 COVID-19 deaths in the past 10 days approximately two weeks later (**APPENDIX TABLE 5**).

Initiating a higher contact reduction earlier is associated with lower peak ICU and ward bed occupancies, even when the threshold to begin increasing contact reductions is the same (**FIGURE 2**). In the scenarios maintaining between a 15% and 20% contact reduction compared to pre-pandemic levels of daily contacts (i.e., similar to Step 3), peak ICU occupancy was between 26 and 34 beds and peak ward bed occupancy was between 78 and 102 beds. Increasing the level of general population contact reduction to between 25% and 27%, led to both a later need for restrictive policies and lower peak hospitalizations. Increasing the level of contact reduction to 30%, compared to pre-pandemic levels of daily contacts, prevented the need for more restrictive policies. The scenario in which contacts were reduced 30% was the only scenario to mitigate the 4^th^ wave such that peak hospital occupancy remained below the peak of the 3^rd^ wave.

### Age distribution of infections and hospitalizations

Compared to previous pandemic waves, children under 12 years compose a greater number and fraction of projected infections and hospitalizations (**FIGURE 3**). In scenarios with and without dynamic behaviour and policy change, children under 12 years represent between 24% and 31% of projected infections between August 1 and December 31, 2021. Infected children are less likely to develop severe disease and are less likely to be hospitalized. As a result, they represent between 6.0% and 8.5% of ward hospitalizations and 3.2% and 4.6% of ICU hospitalizations. Despite their relatively low rate of severe disease requiring hospitalization, without policy intervention to reduce contacts in the community, the projected peak pediatric ICU occupancy is 3.4, 2.6, and 1.4 for scenarios with 15%, 20%, and 25% contact reduction, respectively (compared to a total capacity of 12 pediatric ICU beds for COVID-19 and non-COVID-19 patients) (**APPENDIX TABLE 5**).

**Figure 3.**
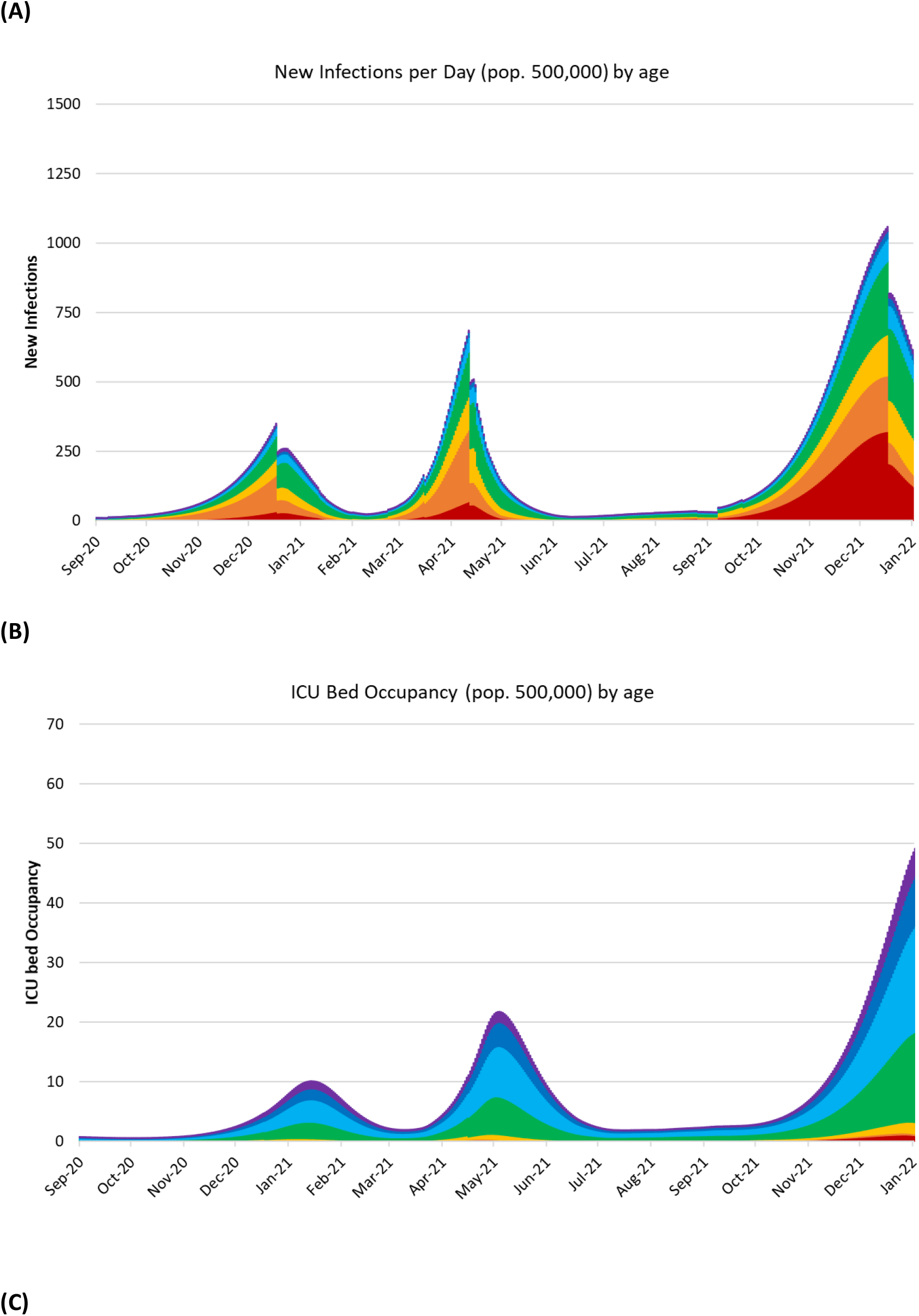
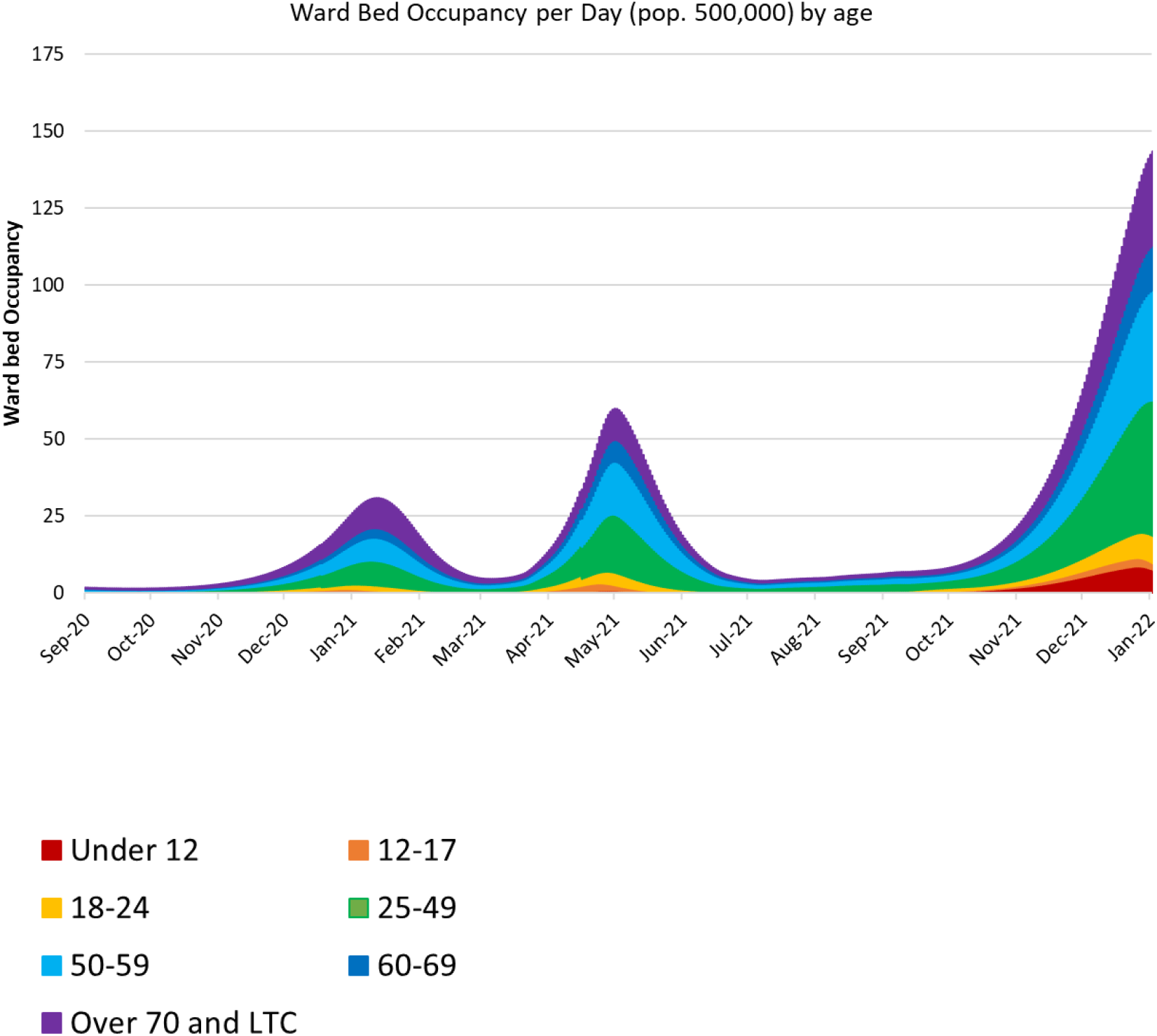
Modelled age distribution of (A) new infections per day, (B) ICU occupancy, and (C) ward bed occupancy for September 2020 through December 2021 in London-Middlesex in and from members of the local community. The scenario shown assumed a 25% contact reduction and no dynamic behaviour change or policy change.

University and college students are mandated to be vaccinated [50], and so we assume that 98% of the student population will be vaccinated. Even so, in scenarios with low levels of contact reduction, 15%, 20%, and 25%, we estimate that 23%, 14%, and 5.4% of the student population, respectively, would become infected over the term without additional mitigation. In scenarios with higher levels of contact reduction, 27% and 30%, and in all cases in which public health measures are reinstituted, the proportion of students infected over the term is less than 4%. Consistent across all scenarios is the connection between on-campus infections and increased community transmission with, on average, each infection in a post-secondary student leading to 0.2 secondary infections in the off-campus community.

### Timing of higher-intensity policy restrictions

Immediately increasing the intensity of policy restrictions to a moderate level of contact reduction, i.e., 27.5% to 30% compared to pre-pandemic levels, can prevent the further need to re-instate high-intensity policies intended to reduce illness and protect hospital resources.

At current levels of contact reduction, re-instatement of high-intensity policy restrictions, such as a stay-at-home order, will be required in the fall of 2021. The timing at which higher-intensity contact reduction efforts begin affects COVID-19 community outcomes. Conditional on the initial level of contact reduction, initiating higher-intensity policies earlier rather than later lowers peak hospital occupancy (**FIGURE 4**). Importantly, the same set of trigger thresholds leads to greater peak occupancy at lower levels of initial contact reduction. Because hospital occupancy information lags infections, at lower initial levels of contact reduction (e.g., 15% to 20%), there are more infections already in the community when the threshold is achieved, leading to higher levels of peak occupancy (**FIGURE 4** and **APPENDIX TABLE 5**).

**Figure 4.**
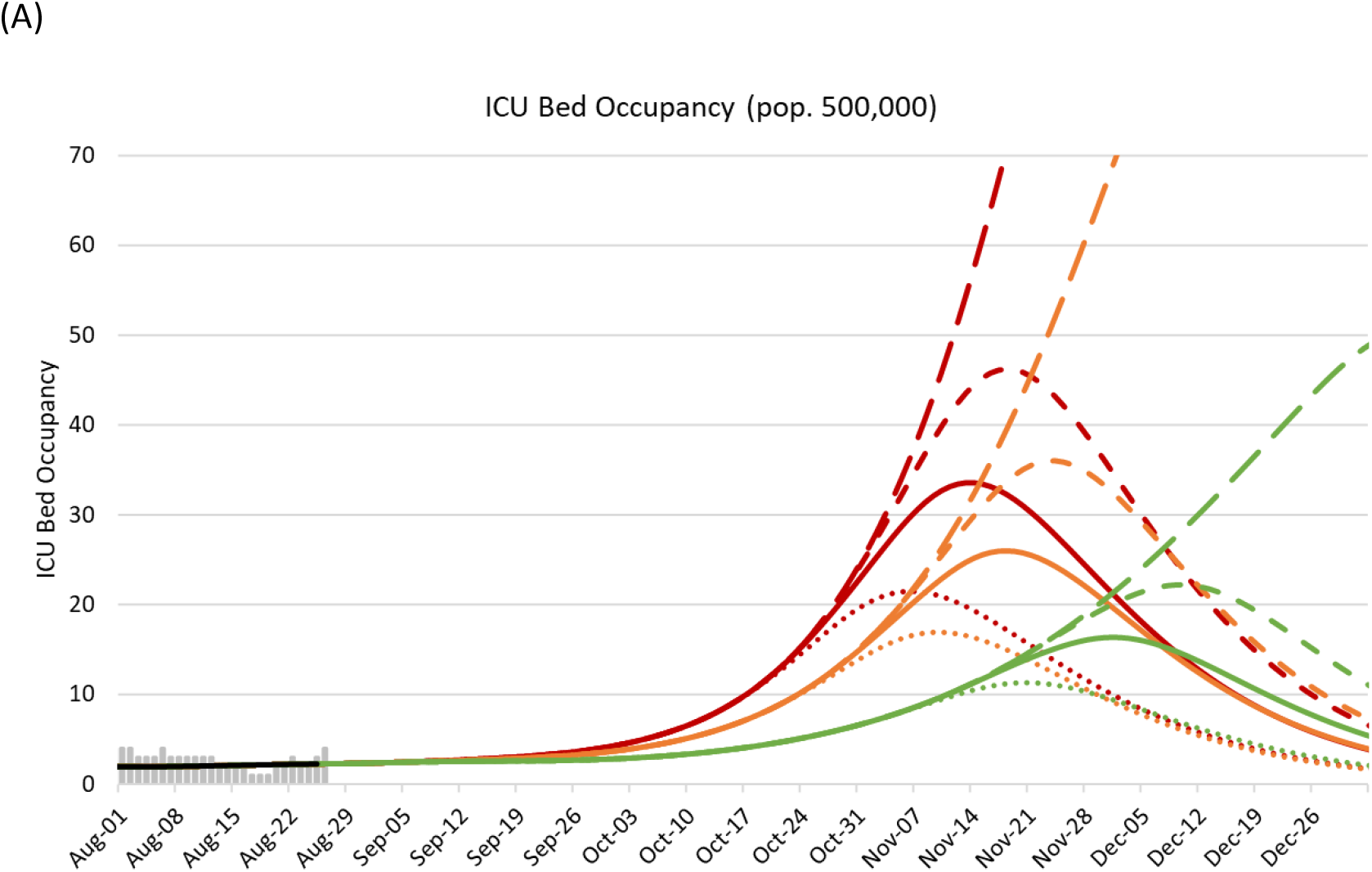
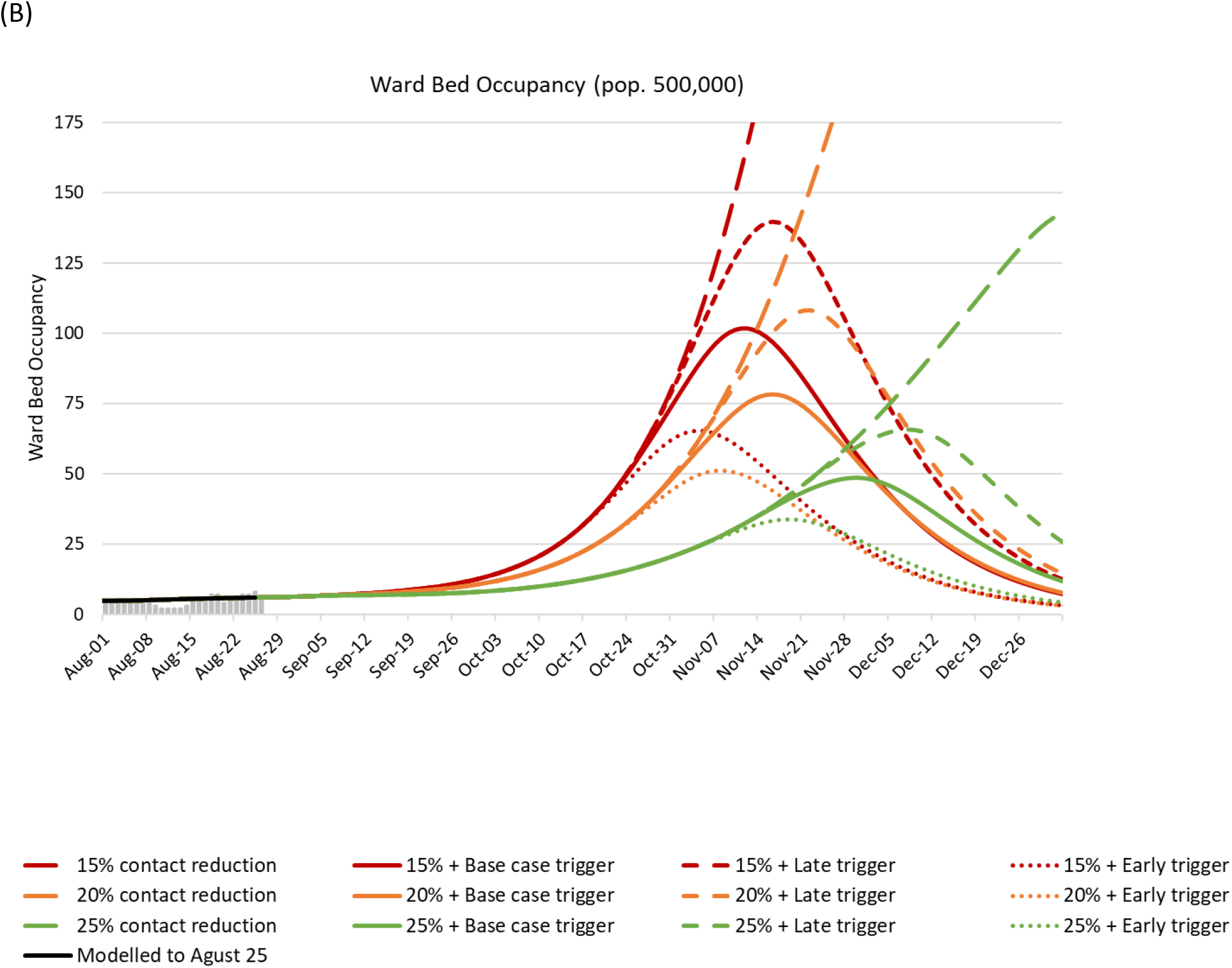
Modelled age distribution of (A) ICU occupancy, and (B) ward bed occupancy for September 2020 through December 2021 in London-Middlesex in and from members of the local community. The scenario shown vary in the level of contact reduction at baseline (15% red; 20% orange; 25% green) and the thresholds used for dynamic behaviour and policy changes: no response, long-dashed line (shown for reference); later response thresholds (10 ICU beds and 7 COVID-19 deaths in 10 days, short-dash line); base case response thresholds (7 ICU beds and 5 COVID-19 deaths in 10 days, solid line); and, earlier response thresholds (5 ICU beds and 3 COVID-19 deaths in 10 days).

### Sensitivity analysis

Many of our model parameters, in particular those that are specific to the Delta variant, are highly uncertain warranting extensive sensitivity analysis. We highlight those that meaningfully impact the policy analysis.

#### Vaccine effectiveness against infection by the Delta variant

The relative performance of vaccines in preventing infection by the Delta variant has been empirically measured in only a few studies to date [18, 44, 46, 51]. In our base case analysis, we assumed that a fully vaccinated person is 90% less likely to be infected by the Alpha variant than an unvaccinated person and that vaccines provide 15% less, i.e., 76.5%, protection against infection with the Delta variant. In sensitivity analysis we considered scenarios in which vaccines provide 80% and 85% protection against infection by the Delta variant. Higher vaccine efficacy is associated with lower numbers of infections and a delayed need for high-intensity policy restrictions. However, even with the most optimistic vaccine effectiveness against the Delta variant, completely averting the need for high-intensity policy restrictions was only possible in scenarios in which the initial level of contact reduction exceeded 20% (**APPENDIX FIGURE 4**).

#### Projected vaccine uptake

In the base case, we assumed observed vaccine uptake until the week of August 15, and then projected forward based on a slowing weekly rate of uptake (**APPENDIX FIGURE 2**). In sensitivity analysis, we considered alternative projections in which at least 85%, 90%, or 95% were vaccinated in each vaccine-eligible age group. Even 95% vaccination in all eligible age groups did not prevent the necessity of high-intensity restrictions from being re-imposed without initial contact reductions exceeding 27% (**APPENDIX FIGURE 6**).

#### Pediatric length of stay

Until recently, pediatric patients were a small fraction of all hospitalized patients and so there is greater uncertainty around their hospital length of stay and outcomes. In the base case, we estimated pediatric hospital and ICU length of stay based on moderately sized patient cohorts [52-54]. In sensitivity analysis we considered a +1 day and +2 day increase to both pediatric ward and critical care hospitalizations. Given the small absolute number of pediatric critical care beds available in our Centre (12 beds) and the true catchment area of the pediatric hospital being four times greater than the modelled population, longer length of stay has the ability to meaningfully impact peak occupancy (approximately a 33% increase) (**APPENDIX TABLE 6**).

## DISCUSSION

With increased transmissibility, increased severity, and potentially lower vaccine protection against infection, the Delta variant has changed the nature of the pandemic [7, 11-16, 23, 44]. It is no longer conceivable to rely on high rates of vaccination in people over the age of 12 years to prevent an overwhelming resurgence of COVID-19 in the Fall of 2021. Our analysis found that at high levels of vaccination and relatively high levels of mask wearing, a moderate additional effort to reduce contacts is necessary to avoid re-instating intensive public health measures. Maintaining contact reductions at the current level, estimated to be a 17% reduction compared to pre-pandemic contact levels, results in exceeding 20% of pre-pandemic ICU capacity by late October, requiring cancellation of elective surgeries. Increasing the contact reduction level to 25% results in exceeding 20% of ICU capacity in the third week of November. Reducing contacts by 27% to 30%, compared to pre-pandemic contact levels, prevents high case volumes and the re-instatement of more restrictive public health measures.

Highly effective vaccination has fundamentally decreased the severity of disease and provides substantial protection against infection, markedly slowing disease transmission [17-21, 46, 47, 51, 55]. Vaccination rates across Canada are among the highest coverage rates in the world [56]. While still highly effective at preventing severe disease against the Delta variant, further increasing vaccination rates among the vaccine-eligible population is not sufficient to mitigate a fourth wave. Policies to achieve an overall 30% contact reduction include maintaining firm limits on indoor gathering size, facilitating work from home, and providing paid sick leave to support quarantine and isolation. Although not specifically considered in our analysis, other measures that have been proposed include frequent testing in school settings and workplaces where physical distancing is not possible [57-62].

Our projections are consistent with other modelling projections. For example, without additional contact reduction and without responsive behaviour change, we estimate 50% to 60% of children under 17 years will become infected over the fall term, consistent with an analysis of school-based transmission that estimated 50% of susceptible children would become infected at school over the fall term with universal masking and low-levels (30%) of vaccine protection [63]. We estimate that a 25% contact reduction leads to approximately 5.4% of the post-secondary student population becoming infected over the term, even with a 98% vaccination rate, consistent with the projection from a model of COVID-19 transmission on college campuses [61]. In addition, three models providing population-level projections for the fall of 2021 in Canada, two at the national level and another for British Columbia, demonstrate a similar relative increase in the number of infections per day compared to the 3^rd^ wave as well as a similar shift in the age distribution of infections towards younger people [64-66].

In-person education is an essential priority for the overall wellbeing, psychosocial development, and educational development of children [1, 67, 68], but it also increases the absolute number of contacts for children. School-based policies such as student cohorts, small class sizes, improved ventilation, routine testing, and vaccine mandates for teachers and staff can reduce in-school transmission [68, 69]. Even so, schools will remain higher contact environments where, particularly in elementary schools where children are unvaccinated, significant outbreaks are possible [69-71]. Ultimately, in order for children to return to in-person education and minimize the impact of increased contacts on community transmission, adults in the community will need to further reduce their contacts.

Our study has several limitations. First, in the Canadian healthcare system, no region is entirely self-contained. During wave 3, patients were transferred for ICU and hospital care between cities, regions, and provinces. In addition, pediatric hospital resources are organized differently from adult resources, the catchment area for the former being approximately three times the size of the latter. Second, our triggers for behaviour and policy change were set around total critical care occupancy, driven largely by adult use; at a 30% contact reduction, peak pediatric ICU demand was 0.07 per 100,000 population (representing 1.4 pediatric ICU beds adjusting for the catchment population of 2 million for the pediatric hospital), which still exceeds 10% of capacity. Higher rates of infection in children may lead to more rapid adoption of behaviour change, but the lower relative capacity in pediatric critical care will require policy makers to respond to rates of utilization in pediatric facilities specifically. Third, our model does not incorporate waning vaccine efficacy. While 65% of the vaccinated population was vaccinated after April 1 (within the past 4 months), our model may underestimate infection risk and disease severity in the older adult population because 75% of people over the age of 70 years and almost all long-term care residents were vaccinated more than 4 months ago [2, 3]. Finally, we do not consider seasonality or a transition towards indoor contacts as the weather gets colder. In all these regards, our projections can be considered conservative.

The *R*_*0*_ of the Delta variant is between 6.75 and 8.0. Notwithstanding high rates of vaccine coverage among the vaccine-eligible (76% fully vaccinated and 83% having at least first dose [3]), approximately 15% of the total population remains ineligible for vaccination heading into the fall of 2021 because they are under 12 years of age. Ongoing mask wearing and reducing contacts by, on average, 30% is required to prevent re-instatement of restrictive policies as hospital resources, again, become overwhelmed. Our analysis indicates that immediate moderate public health measures can prevent the necessity for more intense, disruptive, and difficult to sustain measures later.

## Data Availability

Data used for the development of this model are publicly available.

