## Supplemental Material for "MITIGATING THE 4^th^ WAVE OF THE COVID-19 PANDEMIC IN ONTARIO"

Cipriano LE, Haddara WMR, Sander B

September 1, 2021

**Supplemental Material**

### SUPPLEMENTAL METHODS

#### OVERVIEW

We extended and updated a previously published deterministic dynamic compartmental model of SARS-CoV-2 transmission that predicts health outcomes and resource use in a representative mid-sized city with a full-time population of 500,000 and an additional part-time academic year population of 50,000 post-secondary students [1]. We use the model to project how COVID-19 risk and prevention behaviours of the general population, the return of in-person elementary and secondary schools, and the return of in-person post-secondary education impacts community health outcomes in the Fall of 2021. We consider the impact of several interventions that decrease the probability of disease transmission, including reducing the number of contacts and vaccine coverage, on the number of infected individuals and acute care demand between August 1, 2021 and December 31, 2021.

The model was implemented in Microsoft Excel in Microsoft Office 365. Institutional ethics review was not required for this modeling study.

In these Supplemental Methods, we provide additional detail on the structural features of the model as well as the identification and selection of input parameter values.

### DISEASE SEVERITY AND ACUTE CARE RESOURCE NEED

Disease severity affects the likelihood of case identification through symptom-based detection and the likelihood of being hospitalized and needing specialized health care resources. Asymptomatic infections, representing 31% of infections at all age groups [2], are less likely to be identified and self-isolate. We estimated the age-specific probability of hospitalization and the age-specific fraction of hospitalizations leading to critical care for the original strain of SARS-CoV-2 present in North America using the number of reported diagnosed cases, hospitalizations, and critical care hospitalizations reported in the COVID-19 daily epidemiologic update for Canada, adjusted for the rate of underdiagnosis estimated using serology analysis performed earlier in the pandemic (**APPENDIX TABLE 1**) [3, 4]. We estimated the proportion of hospitalizations leading to critical care for individuals living in long term care (LTC) using the Public Health Agency of Canada dataset which includes information about LTC residency [5].

We assumed the overall probability of hospitalization is 1.63-times higher for infection with the Alpha variant, compared to the original strain dominant in North America in the spring of 2020 [6-8]. We assumed the probability of hospitalization is an additional 1.85-times higher for infection with Delta variant compared to Alpha variant [8-10].

Among critical care patients, we estimated the age-specific probability of requiring respiratory support or renal replacement therapy (RRT) based on the UK Intensive Care National Audit and Research Centre (ICNARC) report describing the care and outcomes of 26,550 critical care COVID-19 patients in the UK [11]. Specifically, we assumed 11.3%, 15.9%, and 14.1% of critical care patients aged 18-49 years, 50-69 years, and 70 years and older require RRT in ICU, and the remainder require respiratory support only [11].

Of pediatric ICU patients in the UK, approximately half (49%) required invasive ventilation, approximately half required continuous vasoactive infusion (47%), and few required RRT (3%) or ECMO (<1%) [12]. So, instead of dividing ICU patients into respiratory support only or RRT as we did with adults, we stratified the pediatric ICU population into respiratory support or vasoactive infusion [12].

**Appendix Table 1. Probability of hospitalization and proportion of hospitalized patients receiving critical care services by age group**

| | Age | | | | | | | |
|---|---|---|---|---|---|---|---|---|
| | **< 18 y** | **18-24 y** | **25-49 y** | **50-59 y** | **60-69 y** | **70+ y** | **LTC** | **Reference** |
| **Probability of hospitalization\|infection - Original strain in North America** | 0.19% | 0.73% | 1.77% | 3.87% | 5.40% | 12.23% | 12.6% | [3-5] |
| **Probability of hospitalization\|infection – Alpha variant** | 0.30% | 1.19% | 2.88% | 6.31% | 8.80% | 19.94% | 20.61% | Calculated (× 1.63 [6-8]) |
| **Probability of hospitalization\|infection – Delta variant** | 0.56% | 2.19% | 5.33% | 11.68% | 16.29% | 36.89% | 38.13% | Calculated (× 1.85 [8-10]) |
| **Critical care, as a proportion of hospitalized patients** | 11.6% | 12.5% | 19.1% | 25.6% | 28.3% | 13.7% | 5.1% | [4, 5] |

CLINICAL OUTCOMES AND ACUTE CARE RESOURCE USE

In our forward-looking analysis, assuming a high prevalence of Delta variant infections (95%), we decreased the mean incubation period from 5.6 days [13] to 4.6 days to account for the shorter incubation period of Delta variant [14].

*Length of stay*

We estimated age-specific average length of stay for COVID-19 patients requiring critical care from the UK ICNARC report, UK Paediatric Intensive Care Audit Network report (for patients under 17 years of age), and a US study of over 42,000 COVID-19 hospitalizations (**APPENDIX TABLE 2**) [11, 12, 15]. ICU requiring RRT has a longer length of stay and higher mortality rate than ICU requiring respiratory support. We estimated the average length of these patients (24.5 days) based on UK critical care patient outcomes [11]. We estimated age-specific average length of stay for patients in hospital but not in ICU, including the length of post-discharge from ICU using the US report [15]. This was consistent with length of stay reported in a UK analysis [16].

For pediatric patients, we estimated the average length of stay for continuous vasoactive infusion to be 5.1 days based on a large US pediatric ICU patient series [17]. We calculated the average length of stay for patients receiving respiratory support to be 3.9 days so that the overall length of stay for COVID-19 observed in pediatric ICU patients in the UK of 4.52 days [12, 18].

As necessary, health states were sub-divided into successive states to ensure realistic distributions for the duration of infectiousness and the duration of hospital resource utilization (e.g., gamma distributions instead of exponential distributions) [19, 20].

**Appendix Table 2. Age-specific average length of stay (days) based on hospital resources used.**

| | Age | | | | | | | |
|---|---|---|---|---|---|---|---|---|
| | **<18 y** | **18-24 y** | **25-49 y** | **50-59 y** | **60-69 y** | **70+ y** | **LTC** | **Reference** |
| **Ward bed only** | 4.5 | 4.6 | 4.6 | 7.0 | 7.0 | 8.8 | 8.8 | [15] |
| **Critical Care requiring respiratory support** | 3.9 [12, 18] | 7.6 [15] | 7.6 [15] | 11.9 | 11.9 | 9.8 | 9.8 | [11, 12, 15, 18] |
| **Critical care requiring vasoactive infusion** | 5.1 [17] | NA | NA | NA | NA | NA | NA | [17] |
| **Post-ICU recovery in ward** | 1.4 | 5.7 | 5.7 | 6.7 | 6.7 | 6.3 | 6.3 | [15] |
| **Critical care requiring RRT** | NA | 24.5 | 24.5 | 24.5 | 24.5 | 24.5 | 24.5 | [11] |

*Mortality*

We estimated age-specific critical care mortality rates for adults using the UK ICNARC report [11] and for patients under 18 years of age using the UK Paediatric Intensive Care Audit Network report [12, 18]. For patients in hospital but not in critical care and for LTC residents outside of hospital, we used the Public

Health Agency of Canada dataset to estimate the age-specific mortality [5]. For the general population and university students, we assumed that there was no mortality risk for individuals with asymptomatic infections or infections not requiring hospitalization.

We did not include mortality from causes other than COVID-19 in the model.

**Appendix Table 3. Age-specific probability of death conditional on disease severity and resource use**

| | Age | | | | | | | |
|---|---|---|---|---|---|---|---|---|
| | **<18 y** | **18-24 y** | **25-49 y** | **50-59 y** | **60-69 y** | **70+ y** | **LTC** | **Reference** |
| **Probability of mortality by disease severity and hospital resource use** | | | | | | | | |
| Asymptomatic | 0% | 0% | 0% | 0% | 0% | 0% | 0% | Assumed |
| Symptomatic but not hospitalized | 0% | 0% | 0% | 0% | 0% | 0% | 20.1% [5] | Assumed |
| Ward only | 0.21% [18] | 0.66% [4] | 1.06% [4] | 3.2% [5] | 7.7% [5] | 26.8% [5] | 50.1% [5] | |
| Critical Care requiring respiratory support | 3.8% [12, 18] | 7.3% [11] | 12.2% [11] | 19.7% [11] | 38.7% [11] | 53.9% [11] | 73.3% [5] | |
| Critical Care requiring RRT | NA | 53.8% | 53.8% | 69.0% | 69.0% | 79.5% | 79.5% | [11] |
| Critical care requiring vasoactive infusion | 3.8% | NA | NA | NA | NA | NA | NA | [12, 18] |
| **Overall calculated mortality rates and reported comparators** | | | | | | | | |
| Model, probability of death conditional on hospitalization | 0.63% | 2.2% | 4.1% | 9.4% | 17.8% | 31.0% | 51.3% | Calculated based on inputs |
| Reported probability of death for hospitalized cases | 1.1% [4] | 2.2% [4] | 4.1% [4] | 9.1% [4] | 19.3% [4] | 30.2% [5] | 51.3% [5] | [4, 5] |
| Model, overall probability of death conditional on infection | 0.003% | 0.05% | 0.2% | 1.1% | 2.8% | 11.2% | 31.8% | Calculated based on inputs |

*Recovery*

In the model, people who survive infection move to a recovered state. People with asymptomatic or mild and moderate infections who are not diagnosed during their infectious period continue to participate in physical distancing consistent with rates in the susceptible population as they will continue to adhere to behaviours consistent with individuals who believe themselves not to have been infected.

DIAGNOSIS BY CLINICAL PRESENTATION AND CONTACT TRACING

Assumptions related to the probability of diagnosis in asymptomatic and symptomatic infections are consistent with the assumptions used in our previous analysis [1]. In the base case analysis we assumed the minimum time from symptom onset to diagnosis to be 2.1 days, consistent with the minimum time to self-assess, seek medical attention, and receive diagnostic results [21]. Using the median time to diagnosis based on symptom-based surveillance alone and symptom-based surveillance in combination with contact tracing [22], we estimated that symptom-based surveillance and contact tracing results in a daily probability of diagnosis of 15.8% in individuals with symptoms and the daily probability of detection from contact tracing of 4.1% in asymptomatic infections. This combination of assumptions resulted in approximately 22% of infected individuals being identified, consistent with the overall rates of diagnosis implied by preliminary serology data in Ontario [23], and somewhat lower than diagnosis rates implied by a later Canadian-wide serology analysis [3]. In this analysis, we do not consider scenarios of routine testing of people without known exposure.

For LTC residents, we assume that twice daily symptom screening results in a 40% daily probability of detection in symptomatic patients and that contact tracing with access to all resident contacts increases that probability to 52%. Contact tracing results in an 8.2% daily probability of diagnosis in asymptomatic cases. We assume this is increased further by routine testing for COVID-19 every 7 days which is recommended for the staff of LTC facilities in Ontario [24]. We assumed this routine testing would be performed using nasopharyngeal swab and PCR analysis with a test sensitivity of 72.1% [25, 26].

DISEASE TRANSMISSION

In our previous work, using exponential regression, we empirically estimated the basic reproduction number, $R_0$, for the original strain circulating in North America to be 3.0 based on Ontario's reported cases between March 7 to March 22 [1, 27]. Using an average duration of infectiousness of 10 days and an average number of close contacts per person of 12.6 [28], we calculate the probability of transmission between a susceptible and an infected person, in the absence of any interventions, to be 0.024 (denoted $\beta_0$ in the equations below).

Using the prevalence of Alpha and Delta variants as reported in the Ontario public health surveillance reports [29], we used a piecewise linear function to estimate the proportion of infections that are the original strain, Alpha variant, and then Delta variant each day (**APPENDIX FIGURE 1**). For dates not yet reported, we projected the trajectory based on the observed trajectories in other Ontario counties. We assumed the Alpha variant to be 1.5-times more infectious than the original strain [30-32] and the Delta variant to be 1.5-times more infectious than Alpha [32, 33]. Therefore, in our analysis, the $R_0$ of the Delta variant is 6.75 [3.0 × 1.5 × 1.5 = 6.75].

**Appendix Figure 1. Proportion of diagnosed infections identified as Alpha variant or Delta variant the Ontario public health surveillance reports [29]**

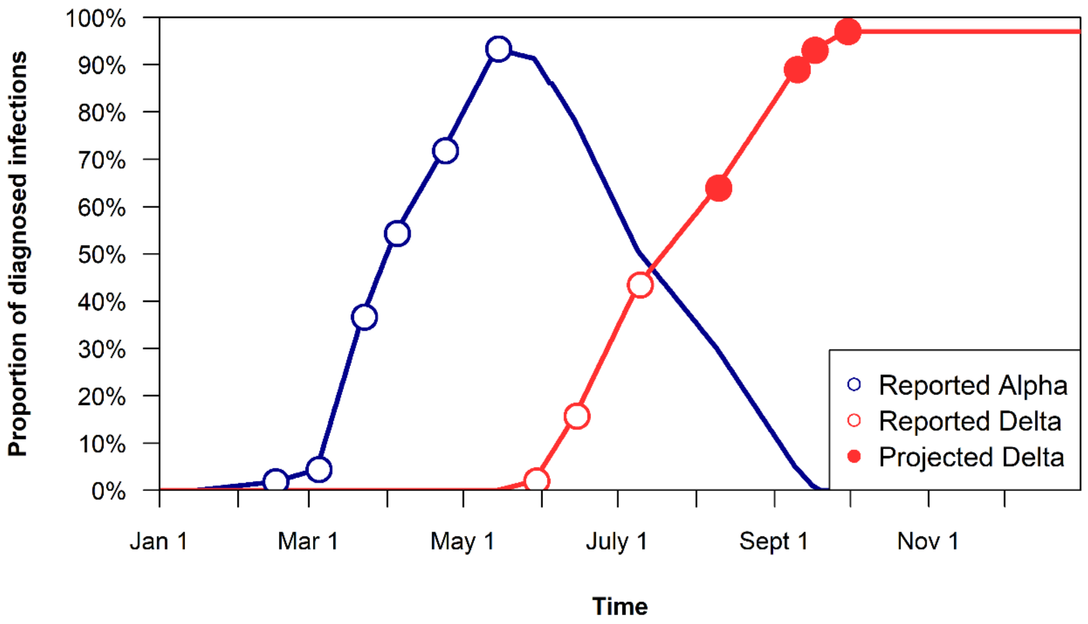


*Daily Number of New Infections*

Each day, we calculated the number of new infections from contacts between susceptible, both vaccinated and unvaccinated, people in Group $i$ subgroup k, $S_{i,k}$, with people in Group $j$, where

$i = j = \{$*0-4 year old, 5-11 year old, 12-17 year old, 18-24 year old, 25-49 year old,*

*50-59 year old, 60-69 year old, 70-plus year old, LTC residents, Post-secondary students*$\}$

and the sub-populations of Susceptible people, $k$, represents time since vaccination or, among the unvaccinated, whether the person is a 'high intensity physical distancer' or a 'low intensity physical distancer' to be

$$\dot{I_{i,j}} = \sum_k S_{i,k}[1-\omega_{i,k}]C_{i,j}[1-\kappa_{i,k}][1-\varepsilon_{ik}]\beta_t \sum_m \frac{I_{j,m}[1-\theta_{j,m}]}{\sum_m N_{j,m}[1-\theta_{j,m}]}$$

where

- $m$ represents subpopulations within Group $j$ with different prevalence, contact behaviours, and mask wearing behaviours

  (i.e., $m = \{$*Unaware of infection status, low-intensity physical distancer; Unaware of infection status, high-intensity physical distancer, Aware Asymptomatic, Aware Symptomatic, Aware Isolation Home, Aware Isolation LTC*$\}$);

- $I_{j,m}$ represents the number of infected people in Group $j$ subgroup $m$;
- $\theta_{j,m}$ represents the relative reduction in transmission due to a reduction in contacts and associated with mask wearing behaviours for people in Group $j$ subgroup $m$;
- $N_{j,m}$ represents the number of individuals in Group $j$ subgroup $m$;
- $C_{ij}$ represents the number of contacts between people in Group $i$ and people in Group $j$;
- $\kappa_{i,k}$ represents the reduction in contacts for people in Group $i$ subgroup $k$;
- $\varepsilon_{i,k}$ represents the reduction in transmission associated with mask wearing behaviours for people in Group $i$ subgroup $k$; and
- $\omega_{i,k}$ represents the reduction in transmission associated with vaccination, including the time since vaccination, for Group $i$ subgroup $k$.

$\beta_t$ represents the probability of transmission conditional on contact between a susceptible and an infected individual at time $t$, adjusting for the mix of variants at each time t, calculated as

$$\beta_t = \beta_0([1 - \alpha - \delta] + \ \alpha_t RR_\alpha + \delta_t RR_\delta)$$

where $\alpha_t$ represents the proportion of Alpha variant infections and $\delta_t$ represents the proportion of Delta variant infections such that $\alpha_t + \delta_t < 1$, and $RR_\alpha$ and $RR_\delta$, respectively, represent the relative risk of transmission if the infected person is infected with the Alpha or Delta variant.

In the model, consistent with our prior work [1], we assume a 90% reduction in contacts for people who are aware of their infection status and in home isolation, which is at the upper end of observed adherence to quarantine instructions in past epidemics [34, 35]. We assume isolation of hospitalized patients is 100% effective at preventing transmission to others.

CONTACT MIXING PATTERNS (PRE-PANDEMIC NORMS)

**General population**: We estimated the average number of close contacts by age for people of each age group using an extrapolation of the 2008 POLYMOD study in Europe to reflect the network structure of the Canadian population [28]. The POLYMOD studies did not include surveys of individuals living in LTC. Similarly, the POLYMOD contact matrices did not include the higher rate of contacts with adult students in a college or university city where these students are more prevalent. We calculated the average number of daily contacts that a person in the general population has with an adult student or a LTC resident by ensuring that the final contact matrix was balanced accounting for contacts with these groups.

**School-based contacts:** We assume that children aged 5-11 years and 12-17 years return to elementary school and secondary school, respectively, on September 7th, 2021. While school is in session, we assume children age 5-11 years have an incremental 8.5 close contacts with other children in the same age group and 0.264 close contacts with an adult (proportionally distributed across adults aged 25-59) based on a French study using RFID technology to measure direct contacts (within 1.5 meters for >5 minutes) in elementary schools [36]. Using a similar study performed in a U.S. high school using a wireless sensor technology identifying contacts within 3 meters for >5 minutes, we assume teens aged 12-17 years have an incremental 23 close contacts with other teenagers in the same age group and 1.30 close contacts with an adult [37]. To calculate these incremental contacts, we subtracted the age-specific school-based contacts already measured in POLYMOD to avoid double counting [28].

**University students**: Consistent with our prior analysis and a fairly consistent literature based on contact surveys in post-secondary students [38-41], we assume that students have 23.7 contacts per day and that 60% of those contacts are with other university students [38]. The remainder of a student's contacts are with members of the 'general population' which includes staff and faculty of the university as well as other members of the community when students are in transit, shopping, and working in jobs in the community, or non-student members of their household based on the age distribution of contacts for 18-24 year olds in the POLYMOD contact matrices.

**Long-term care residents**: Consistent with our prior analysis, we assume that long term care residents have 19.9 resident-resident contacts per day and 13.7 resident-staff contacts per day based on an RFID study of a Canadian long-term care facility [42, 43]. In addition, we include 0.48 visitors per day based on the distribution of visit frequency in the 2012 Ohio Nursing Home Family Satisfaction Survey [44]. Thus, in total, LTC residents have 14.2 contacts with the general population each day. We assumed no direct contact with university students.

### COMMUNITY BEHAVIOUR CHANGES IN RESPONSE TO COVID-19

We divide the population over 12 years of age, including post-secondary students, into two groups: 'High-intensity physical distancers' and 'Lower-intensity physical distancers', initially to capture heterogeneity in adherence to policy whether by the nature of employment, circumstances, or choice. Once fully vaccinated, we assume fully vaccinated individuals join the 'lower-intensity physical distancing' group. We do not stratify children under 12 years of age or LTC residents; they are all assumed to be 'high-intensity physical distancers'. In the analyses presented, low-intensity physical distancers reduce their contacts at 75% the rate of high-intensity physical distancers. Scenarios presented vary in the level of contact reduction in the high-intensity physical distancer group.

**Mask wearing**: Based on an Angus Reid poll of Canadians taken in August of 2020, we assume that high-intensity physical distancers wear masks for 86% of their contacts that low-intensity physical distancers (and fully vaccinated people) wear masks for 38% of their contacts [45]. Therefore, on average in the model prior to widespread vaccination, people over 12 years of age wear a mask for 75% of their contacts. In schools, contacts occur in settings with universal mask requirements and so we assume masks are worn for 86% of contacts for children under 12 years of age (school and out of school combined). In a wide range of studies with different design, masks have been shown to be highly effective at reducing transmission [46, 47]. We assume the effectiveness of cloth masks in reducing disease transmission is 46% based on the reduction in transmission observed in German cities after the introduction of a mask mandate using synthetic control methods as an approach to reduce confounding [48].

**Dynamic behaviour response**: In some scenarios, we consider the impact of dynamic behaviours in response to COVID-19 outcomes in the community. These changes in behaviour are intended to capture both individual decision making, as observed in a U.S. study of responsive protective behaviours in the absence of government intervention [49], and coordinated policy changes that may be instituted in the community. To inform relevant triggers for policy changes, we relied on locally relevant thresholds and the observed timing of policy interventions in Waves 2 and 3 to identify that substantial reductions in access to other health care services occurs if 15 critical care beds (about 20% of normal critical care capacity in a city of 500,000 [50]) were occupied by COVID-19 patients.

In our analysis, we considered two threshold levels triggering an increase in contact reductions. In the base case, we assumed that the level of contact reduction increases by 0.75% per day if the number of COVID-19 patients in critical care exceeds 7 (about 10% of normal critical care capacity) and by an additional 1.5% each day if the number of COVID-19 deaths in the past 10 days exceeds 5. In alternative scenarios, we used lower and higher thresholds. In the lower-threshold scenario, we assumed that the level of contact reduction increases by 0.75% per day if the number of COVID-19 patients in critical care exceeds 5 and by an additional 1.5% each day if the number of COVID-19 deaths in the past 10 days exceeds 3. In the higher-threshold scenario, we assumed that the level of contact reduction increases by 0.75% per day if the number of COVID-19 patients in critical care exceeds 10 and by an additional 1.5% each day if the number of COVID-19 deaths in the past 10 days exceeds 7. We assumed a maximum level of contact reduction of 50% in adults and 70% in children and teenagers, representing, on the higher end of this range, the closure of schools.

**Contact Reduction and Mask Wearing in Long term care**: We assume that LTC residents consistently have a 50% reduction in their average contacts (from 34.1 contacts to 17.05 contacts) compared to pre-pandemic normal contact patterns. We further assume that of their remaining contacts, 86% are protected by cloth masks providing an overall 46% reduction in disease transmission [48]. We do not vary the level of contact reduction or mask wearing in LTC in the scenario analyses.

VACCINATION

*Vaccine Uptake*

We implemented first and second dose vaccine uptake using actual age-specific weekly consumption data published by the local health unit until August 15, 2021 [51]. We projected uptake over the subsequent ten weeks using a slowly decreasing weekly rate of uptake (**APPENDIX FIGURE 2**). Because vaccination is mandated, we assume a 98% vaccination rate among post-secondary students [52].

In scenarios in which we explore higher rates of vaccine uptake, additional first-dose vaccinations occur in the first week of September followed by second-dose vaccinations in the first week of October.

**Appendix Figure 2. Proportion of people in each vaccine-eligible age group to have received (A) one dose and (B) two doses of COVID-19 vaccine. Solid lines represent weekly data provided on the local health unit dashboard [51]. Dashed lines represent projections used in the base case analysis.**

**(A)**

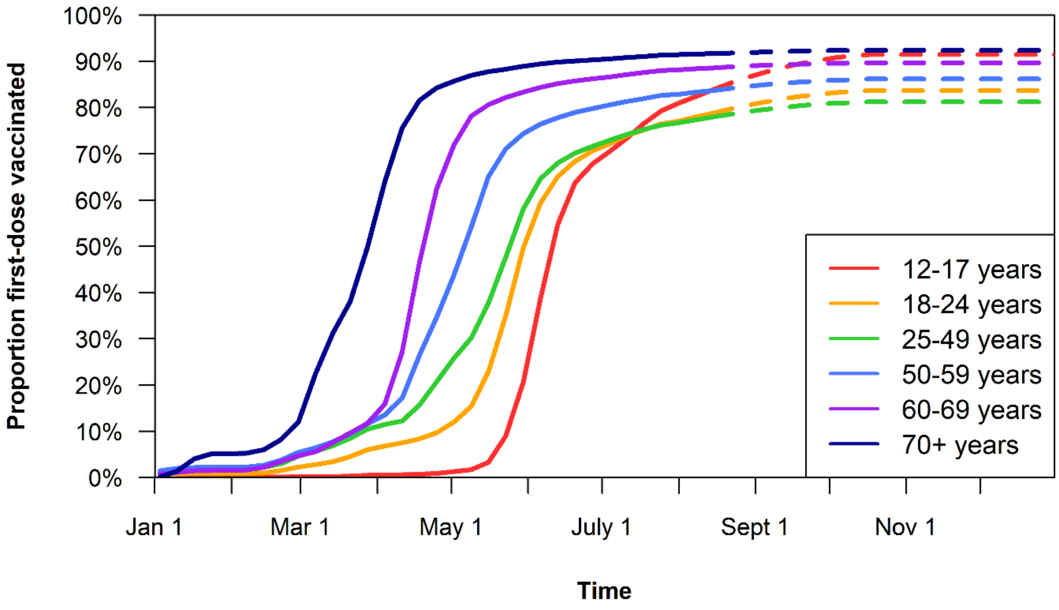


**(B)**

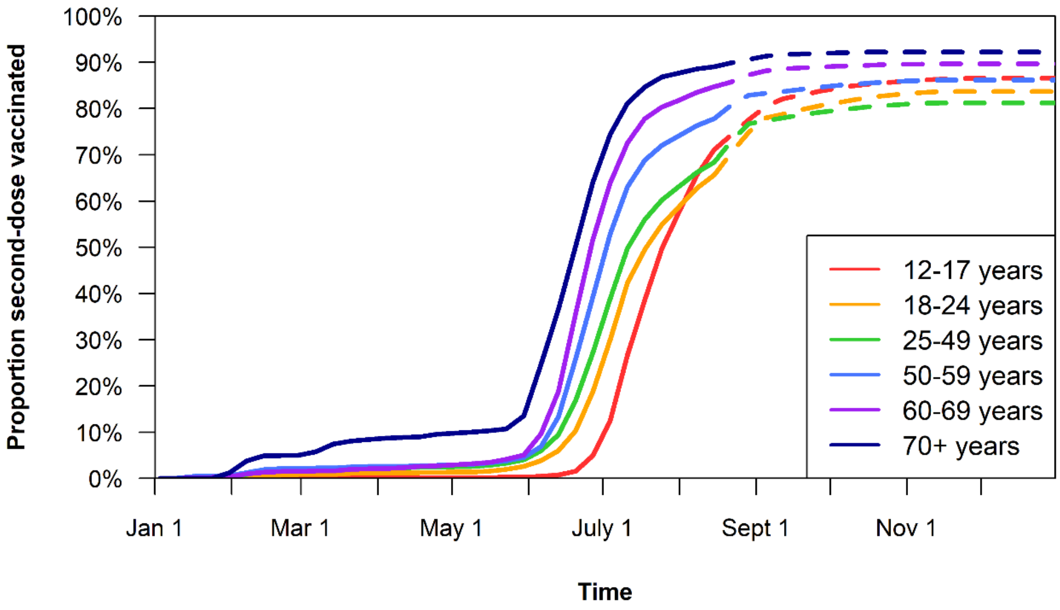

*Vaccine Effectiveness*

We assume efficacy post-first dose and post-second dose takes full effect only two-weeks after receiving the vaccination. Against all variants we assumed the first dose of a two-dose vaccine series provided 58% protection against infection [53-55].

The estimated reduction in infection risk from the Alpha variant conferred by full vaccination ranges from 73% [56] to 97% [57-59]. In the base case, we assumed that full vaccination reduces the probability of infection by the Alpha variant by 90% [54, 60-65]. Fewer studies have evaluated the reduction in infection risk for the Delta variant; one found no difference in the reduction in protection against infection compared to Alpha [53], two found that vaccines provided 15% less protection against infection with Delta [59, 63], and one found 53% less protection against infection with Delta [64]. In the base case, we assumed that full vaccination reduces the probability of infection by Delta by 76.5% (15% less than Alpha) (**APPENDIX TABLE 3**).

Large negative-case control studies and propensity matched control studies have estimated full vaccination reduces the probability of symptomatic infection and the probability of hospitalization [64, 66-68]. To avoid double counting the benefit already conferred from the reduced risk of infection, we calculate the risk reduction for symptomatic disease and the risk reduction of hospitalization conditional on infection. The probability of symptomatic disease is thus reduced by 45% conditional on infection, which in this case leads to a 2.0-fold increase to the probability of asymptomatic infection in vaccinated individuals who become infected. Further, we estimate a 60% reduction in the risk of hospitalization conditional on infection in vaccinated individuals. Conditional on infection, we assume that vaccination provides the same relative reduction in symptomatic infection and hospitalization for both Alpha and Delta variant infections. Collectively, these assumptions lead to vaccination reducing hospitalizations by 96% against the Alpha variant and 91% against the Delta variant, broadly consistent with population level estimates of effectiveness, but substantially higher than the low-end estimates of overall vaccine effectiveness against Delta [55, 63-66]. We calculate overall vaccine effectiveness as:

$$\text{Vaccine overall effectiveness against hospitalization}$$
$$= 1 - (1 - \text{Reduction in Infection}) \times (1 - \text{Reduction in hospitalization conditional on infection})$$

**Appendix Table 4. Reduction in the probability of infection given a contact between a susceptible vaccinated person and an infected person**

| | Days since first dose vaccination | | | | Days since second dose vaccination | | | |
|---|---|---|---|---|---|---|---|---|
| | **0-4** | **5-9** | **10-14** | **14+** | **0-4** | **5-9** | **10-14** | **14+** |
| **Alpha variant** | 0% | 16% | 44% | 58% | 58% | 66% | 74% | 90% |
| **Delta variant** | 0% | 16% | 44% | 58% | 58% | 62% | 67% | 76.5% |
| **Reference** | Assumed | [59] | [59] | [53] | Assumed | Assumed | Assumed | [53-55, 57-64] |

MODEL CALIBRATION

The model has been iteratively calibrated throughout the pandemic to the local conditions in the City of London and Middlesex County in support of local hospital decision making. Notably, we exclude some cases from the end of November and early December from calibration because they were due to a hospital-based outbreak, not community transmission. In preparation for this analysis, calibration to the recovery from the 3rd wave (February to June 2021) allowed us to estimate the current number of infections in the community (in each age group) and the level of contact reduction on August 25, 2021 (**APPENDIX FIGURE 3**). Ultimately, we the estimated level of contact reduction in August 2021, with low-level government restrictions in place (e.g., ‘Step 3’ [69]), to be 17.5% compared to pre-pandemic contact levels.

**Appendix Figure 3. Results of calibration to (A) ICU bed occupancy and (B) ward bed occupancy by local patients over the COVID-19 in the County of Middlesex, Ontario, including the City of London. Notably, we exclude some cases from the end of November and early December from calibration because they were due to a hospital-based outbreak, not community transmission.**

**(A)**

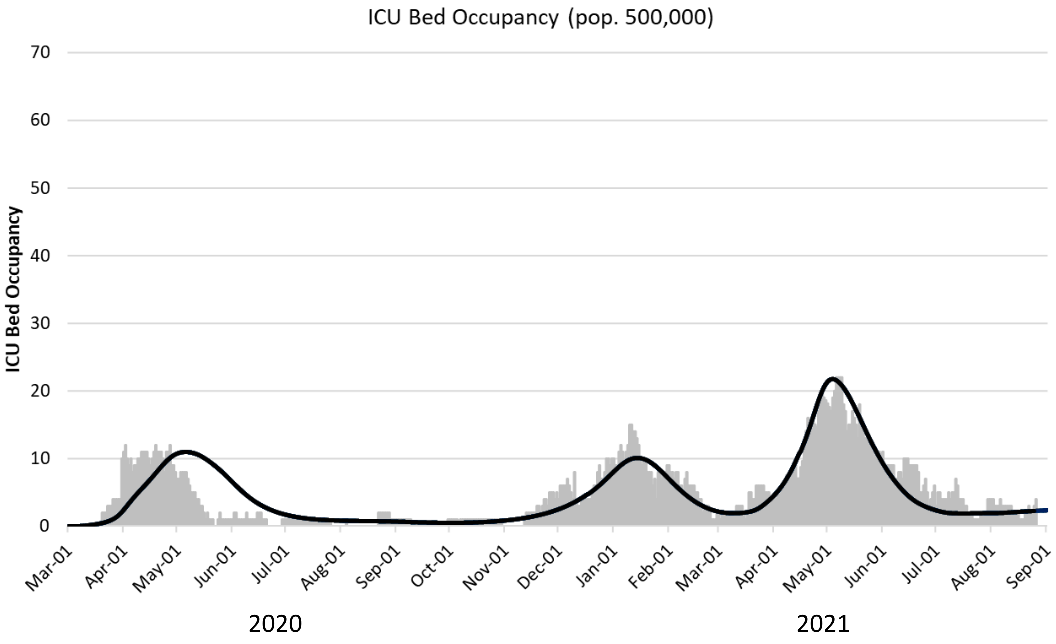


**(B)**

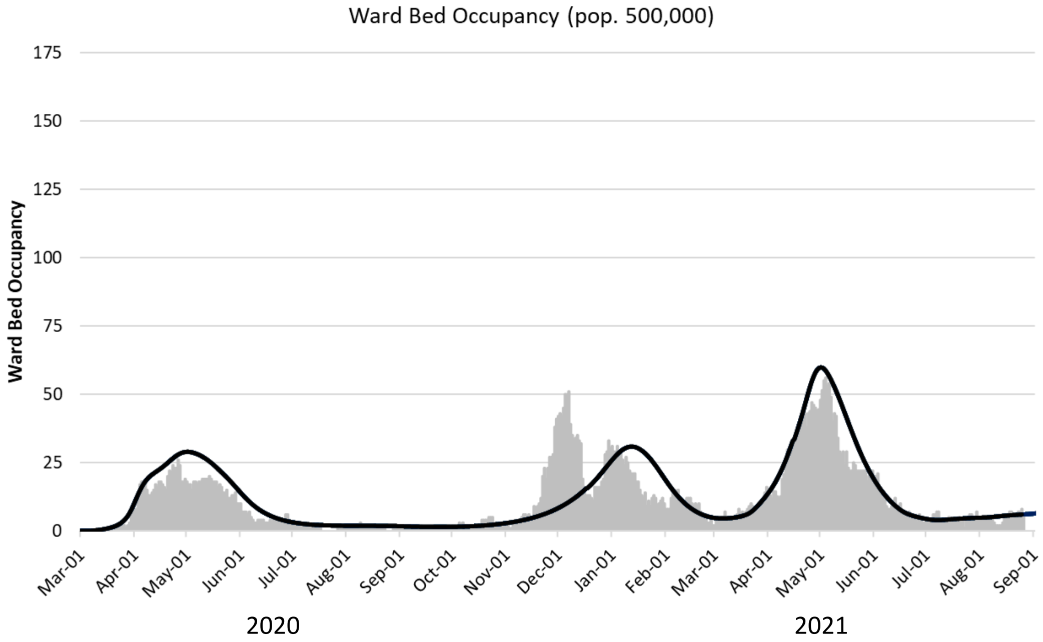

### SUPPLEMENTAL RESULTS

**Appendix Figure 4. Sensitivity analysis on the effectiveness of vaccines to prevent infection with the Delta variant.** Projected (A) new infections per day, (B) ICU occupancy, and (C) ward bed occupancy for August 2021 through December 2021 in London-Middlesex in and from members of the local community. To assist in visualizing the impact of the parameter value on the results, scenarios shown assume no dynamic behaviour change or policy change. Scenarios vary on their level of contact reduction (20%, orange; 25%, green; 27.5%, blue) and the effectiveness of vaccines to prevent infection with the Delta variant (76.5%, base case, solid line; 80%, long-dashed line; 85%, dotted line).

Legend:

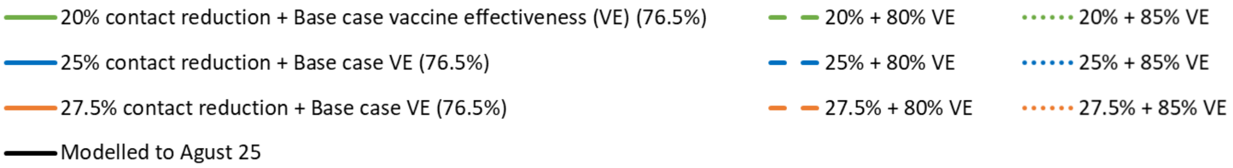


**(A)**

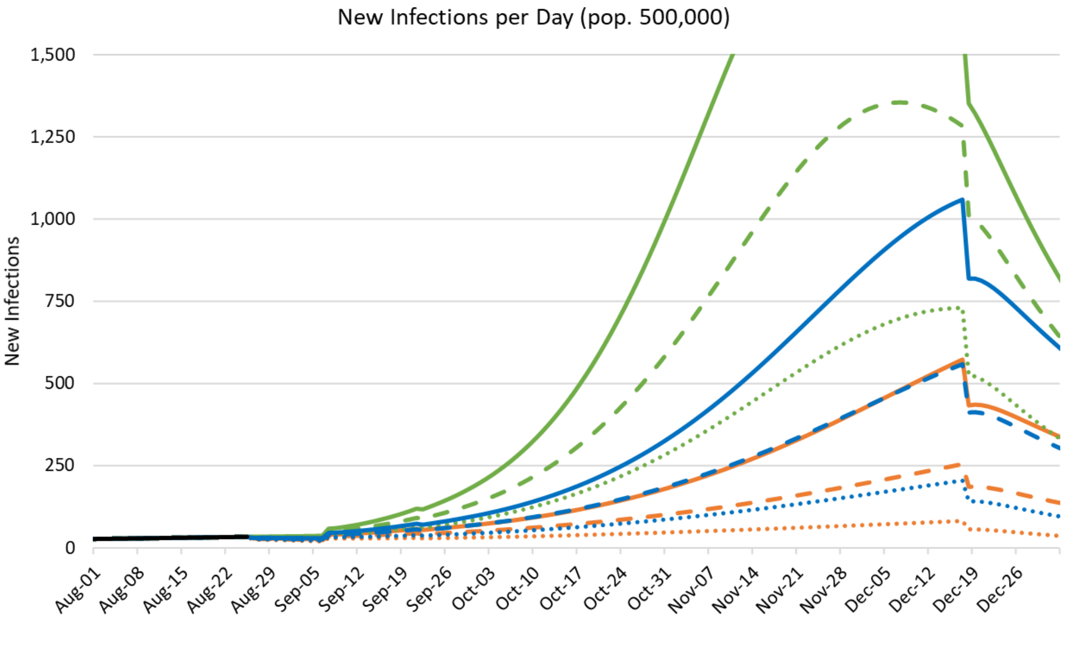

(B)

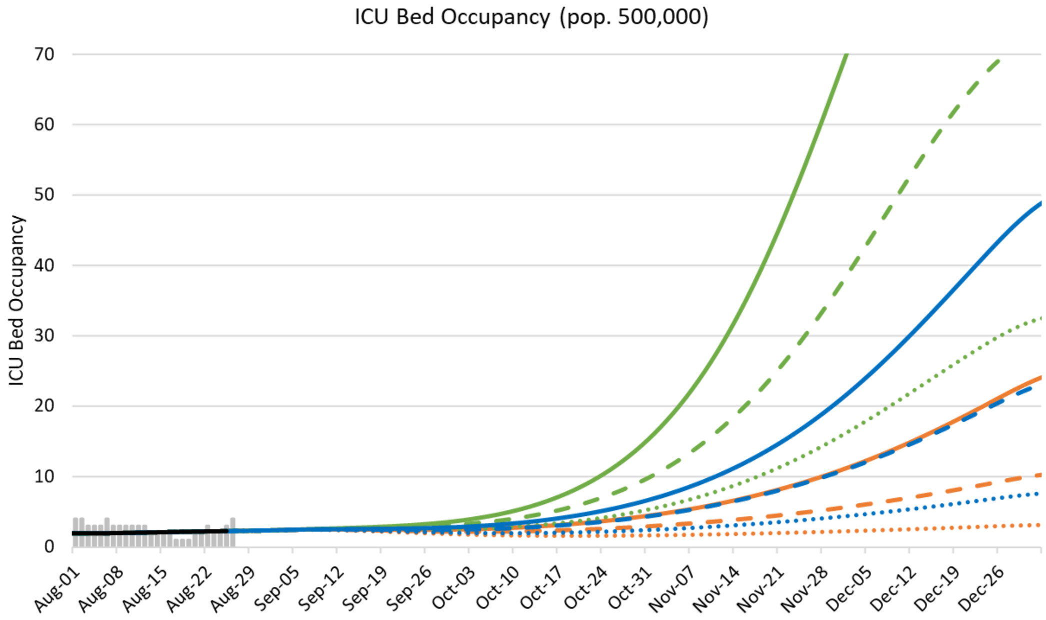

ICU Bed Occupancy (pop. 500,000)
ICU Bed Occupancy
70
60
50
40
30
20
10
0
Aug-01
Aug-08
Aug-15
Aug-22
Aug-29
Sep-05
Sep-12
Sep-19
Sep-26
Oct-03
Oct-10
Oct-17
Oct-24
Oct-31
Nov-07
Nov-14
Nov-21
Nov-28
Dec-05
Dec-12
Dec-19
Dec-26


(C)

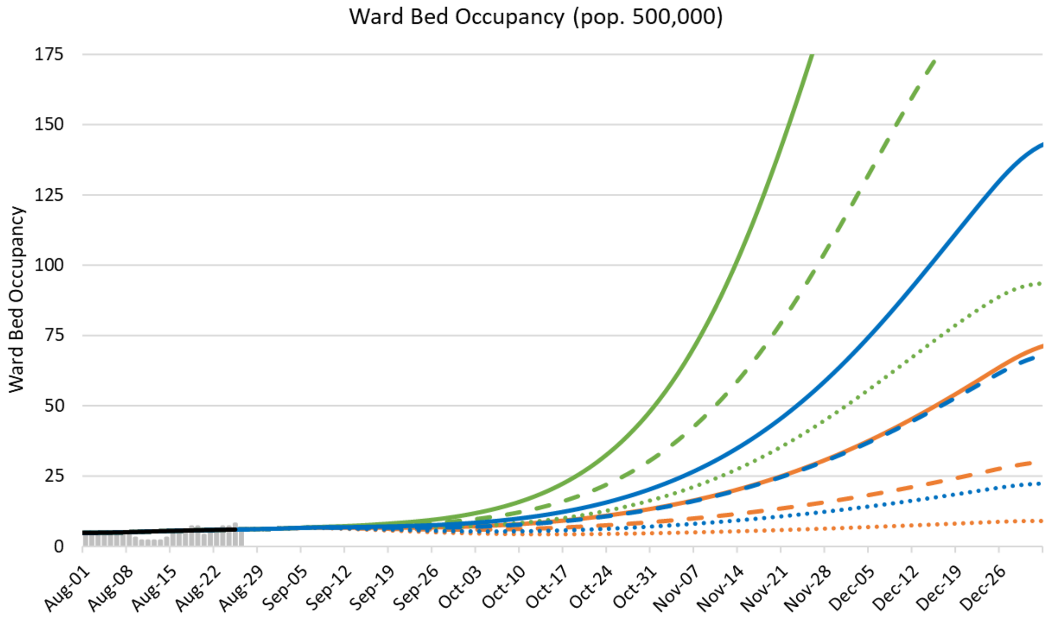

Ward Bed Occupancy (pop. 500,000)
Ward Bed Occupancy
175
150
125
100
75
50
25
0
Aug-01
Aug-08
Aug-15
Aug-22
Aug-29
Sep-05
Sep-12
Sep-19
Sep-26
Oct-03
Oct-10
Oct-17
Oct-24
Oct-31
Nov-07
Nov-14
Nov-21
Nov-28
Dec-05
Dec-12
Dec-19
Dec-26

**Appendix Figure 5. Sensitivity analysis on vaccine uptake.** Projected (A) new infections per day, (B) ICU occupancy, and (C) ward bed occupancy for August 2021 through December 2021 in London-Middlesex in and from members of the local community. To assist in visualizing the impact of the parameter value on the results, scenarios shown assume no dynamic behaviour change or policy change. Scenarios vary in the level of contact reduction (23.5%, orange; 25%, green; 27.5%, light blue; 30%, dark blue) and the minimum level of vaccine coverage achieved in all age groups (85%, solid line; 90%, long-dashed line; 95%, dotted line). Incremental first dose vaccines were implemented the first week of September and second dose vaccines 4-weeks later in the first week of October.

Legend:

- 22.5% contact reduction + 85% vaccine uptake
- 25% contact reduction + 85% vaccine uptake
- 27.5% contact reduction + 85% vaccine uptake
- 30% contact reduction + 85% vaccine uptake
- Modelled to Agust 25
- 22.5% + 90% vaccine uptake
- 25% + 90% vaccine uptake
- 27.5% + 90% vaccine uptake
- 30% + 90% vaccine uptake
- 22.5% + 95% vaccine uptake
- 25% + 95% vaccine uptake
- 27.5% + 95% vaccine uptake
- 30% + 95% vaccine uptake

**(A)**

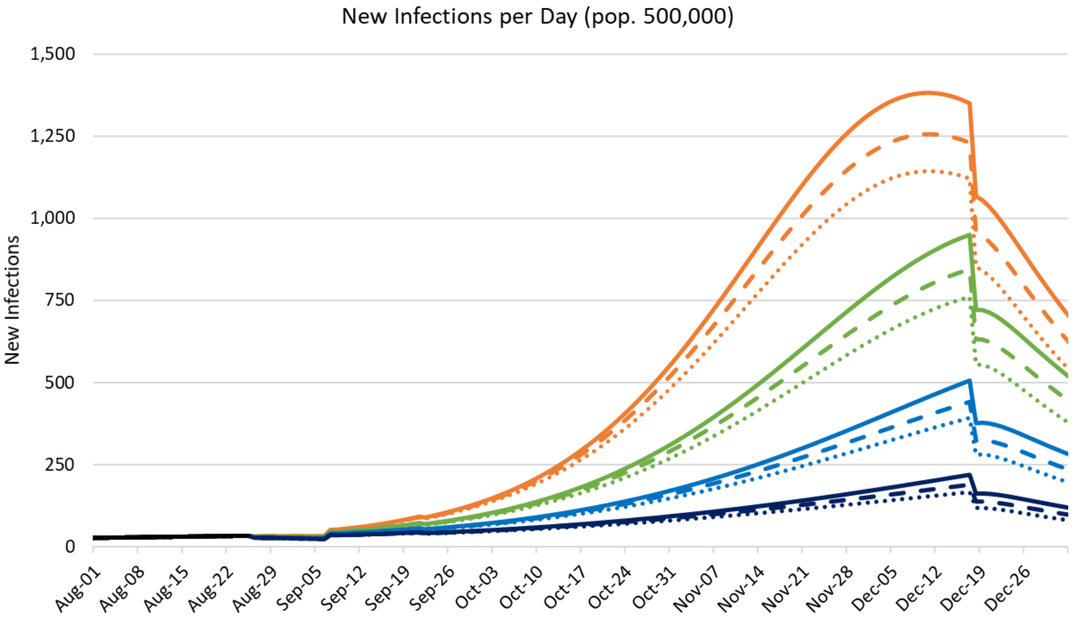

**(B)**

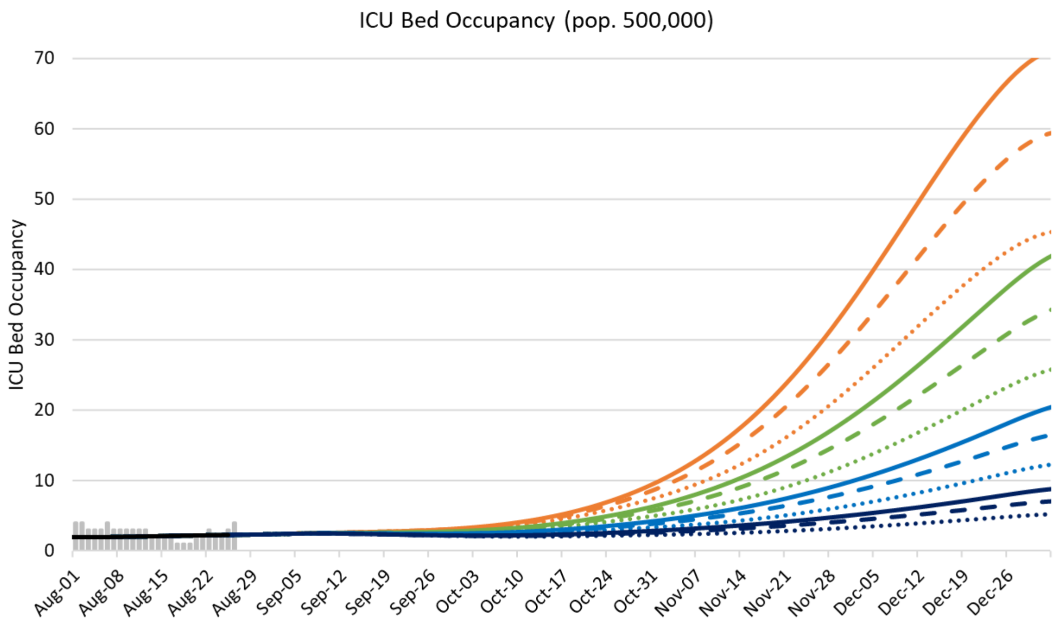
ICU Bed Occupancy (pop. 500,000)
ICU Bed Occupancy
70
60
50
40
30
20
10
0
Aug-01
Aug-08
Aug-15
Aug-22
Aug-29
Sep-05
Sep-12
Sep-19
Sep-26
Oct-03
Oct-10
Oct-17
Oct-24
Oct-31
Nov-07
Nov-14
Nov-21
Nov-28
Dec-05
Dec-12
Dec-19
Dec-26

**(C)**

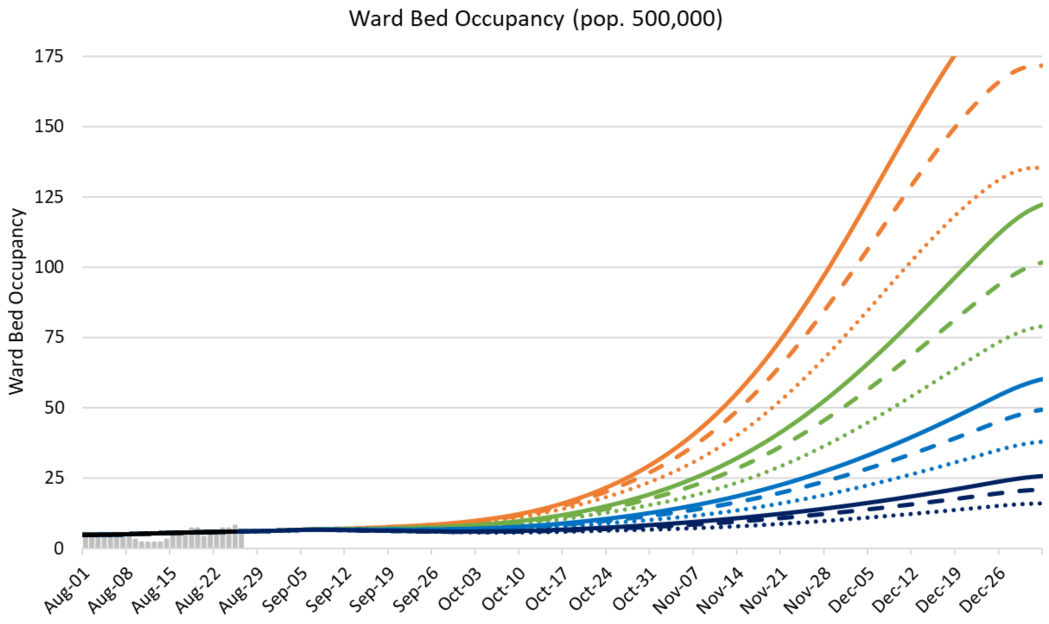
Ward Bed Occupancy (pop. 500,000)
Ward Bed Occupancy
175
150
125
100
75
50
25
0
Aug-01
Aug-08
Aug-15
Aug-22
Aug-29
Sep-05
Sep-12
Sep-19
Sep-26
Oct-03
Oct-10
Oct-17
Oct-24
Oct-31
Nov-07
Nov-14
Nov-21
Nov-28
Dec-05
Dec-12
Dec-19
Dec-26

**Appendix Table 5. COVID-19 community health outcomes for various levels of contact reduction and three different thresholds for initiating high-intensity physical distancing policies and business closures.** In these scenarios, the community begins increasing the level of contact reduction by 0.75% per day after the ICU occupancy threshold is exceeded, and by an additional 1.5% per day after the COVID-19 deaths threshold is exceeded. The maximum level of contact reduction was set to 50% in adults and 70% in children and teens. In the base case, we assumed that contact reduction would begin to increase after 7 ICU beds were occupied with COVID-19 patients (representing 10% of pre-COVID capacity) and, more intensely, when there were 5 COVID-19 deaths within 10 days in the community. In the 'Later' scenario, the triggers were set at 10 occupied ICU beds and 7 deaths in 10 days; in the 'Earlier' scenario, the triggers were set at 5 occupied ICU beds and 3 deaths in 10 days.

| **Contact reduction:** | **15%** | | | | **20%** | | | | **25%** | | | | **30%** |
|---|---|---|---|---|---|---|---|---|---|---|---|---|---|
| | **No trigger** | **Later** | **Base case** | **Earlier** | **No trigger** | **Later** | **Base case** | **Earlier** | **No trigger** | **Later** | **Base case** | **Earlier** | **No trigger** |
| **Trigger activated (month-day)** | | | | | | | | | | | | | |
| ICU Occupancy | | Oct 18 | Oct 12 | Oct 06 | | Oct 25 | Oct 18 | Oct 11 | | Nov 12 | Nov 03 | Oct 25 | |
| Deaths in 10 days | | Oct 31 | Oct 26 | Oct 17 | | Nov 07 | Nov 01 | Oct 22 | | Nov 27 | Nov 18 | Nov 05 | |
| **Number of infections between August 1 and December 31, 2021** | | | | | | | | | | | | | |
| < 12 years | 37,454 | 13,531 | 10,075 | 6,420 | 31,435 | 11,178 | 8,187 | 5,243 | 16,000 | 8,066 | 5,924 | 3,944 | 4,458 |
| 12-17 years | 23,177 | 9,939 | 7,620 | 4,994 | 19,180 | 8,038 | 6,042 | 3,957 | 9,731 | 5,672 | 4,270 | 2,896 | 2,830 |
| 18-50 years | 67,737 | 17,033 | 12,831 | 8,590 | 49,890 | 13,916 | 10,393 | 7,040 | 20,816 | 9,904 | 7,510 | 5,328 | 5,744 |
| > 50 years | 20,787 | 4,920 | 3,639 | 2,365 | 15,294 | 4,052 | 2,975 | 1,962 | 6,219 | 2,886 | 2,156 | 1,497 | 1,658 |
| **Number of hospitalizations between August 1 and December 31, 2021** | | | | | | | | | | | | | |
| Ward | 2,960 | 771 | 576 | 378 | 2,233 | 635 | 470 | 312 | 955 | 454 | 341 | 238 | 261 |
| ICU | 725 | 184 | 137 | 90 | 545 | 152 | 112 | 74 | 230 | 108 | 81 | 57 | 63 |
| Ward, < 18 years | 234 | 89 | 67 | 43 | 196 | 74 | 54 | 35 | 101 | 53 | 39 | 26 | 29 |
| ICU, < 18 years | 31 | 12 | 9 | 6 | 26 | 10 | 7 | 5 | 13 | 7 | 5 | 3 | 4 |
| **Peak hospital occupancy** | | | | | | | | | | | | | |
| Ward | 412 | 140 | 102 | 65 | 316 | 108 | 78 | 51 | 141 | 66 | 49 | 34 | 30 |
| ICU | 146 | 46 | 34 | 21 | 112 | 36 | 26 | 17 | 48 | 22 | 16 | 11 | 10 |
| Ward, < 18 years | 26.8 | 14.4 | 10.9 | 7.2 | 20.9 | 11.0 | 8.2 | 5.5 | 11.4 | 6.5 | 4.9 | 3.5 | 2.8 |
| ICU, < 18 years | 3.4 | 1.8 | 1.4 | 0.9 | 2.6 | 1.4 | 1.0 | 0.7 | 1.4 | 0.8 | 0.6 | 0.4 | 0.4 |

**Appendix Table 6. Sensitivity analysis on pediatric length-of-stay on peak pediatric hospital occupancy.** Base case estimates for pediatric length of stay are presented in **Appendix Table 2.** In the model, there were 100,000 people aged 0-17 (26,000 0-4 years; 39,500 5-11 years; and 34,500 12-17 years). We present results without and with a dynamic behaviour response. In the base case scenario with dynamic response, we assumed that contact reduction would begin to increase after 7 ICU beds were occupied with COVID-19 patients (representing 10% of pre-COVID capacity) and, more intensely, when there were 5 COVID-19 deaths within 10 days in the community.

| | **15% contact reduction** | | | **20% contact reduction** | | | **25% contact reduction** | | | **30% contact reduction** | | |
|---|---|---|---|---|---|---|---|---|---|---|---|---|
| | **Base case** | **+ 1d** | **+ 2d** | **Base case** | **+ 1d** | **+ 2d** | **Base case** | **+ 1d** | **+ 2d** | **Base case** | **+ 1d** | **+ 2d** |
| **Without dynamic behaviour response trigger** | | | | | | | | | | | | |
| **Number between August 1 and December 31, 2021** | | | | | | | | | | | | |
| Infections (% of < 18 year old population) | 60,631 (60.6%) | | | 50,615 (50.6%) | | | 25,731 (25.7%) | | | 7,288 (7.2%) | | |
| Hospitalizations | | | | | | | | | | | | |
| Ward | 234.0 | | | 196.5 | | | 100.9 | | | 28.6 | | |
| ICU | 30.7 | | | 25.8 | | | 13.2 | | | 3.8 | | |
| **Peak occupancy** | | | | | | | | | | | | |
| Ward | 26.8 | 31.1 | 35.4 | 20.9 | 24.3 | 27.8 | 11.4 | 13.2 | 15.0 | 2.8 | 3.2 | 3.7 |
| ICU | 3.4 | 3.9 | 4.5 | 2.6 | 3.1 | 3.5 | 1.4 | 1.7 | 1.9 | 0.35 | 0.41 | 0.47 |
| **With base case dynamic behaviour response trigger** | | | | | | | | | | | | |
| **Number between August 1 and December 31, 2021** | | | | | | | | | | | | |
| Infections (% of < 18 year old population) | 17,694 (17.7%) | | | 14,229 (14.3%) | | | 10,194 (10.2%) | | | 6,997 (7.0%) | | |
| Hospitalizations | | | | | | | | | | | | |
| Ward | 67.3 | | | 54.4 | | | 39.2 | | | 27.4 | | |
| ICU | 8.8 | | | 7.1 | | | 5.1 | | | 3.6 | | |
| **Peak occupancy** | | | | | | | | | | | | |
| Ward | 10.9 | 12.6 | 14.2 | 8.2 | 9.5 | 10.8 | 4.9 | 5.7 | 6.5 | 2.7 | 3.1 | 3.6 |
| ICU | 1.4 | 1.6 | 1.8 | 1.0 | 1.2 | 1.4 | 0.62 | 0.73 | 0.83 | 0.34 | 0.40 | 0.45 |

10. Twohig KA, Nyberg T, Zaidi A, Thelwall S, Sinnathamby MA, Aliabadi S, et al. Hospital admission and emergency care attendance risk for SARS-CoV-2 delta (B.1.617.2) compared with alpha (B.1.1.7) variants of concern: a cohort study. The Lancet Infectious Diseases. doi: 10.1016/S1473-3099(21)00475-8.

11. Intensive Care National Audit & Research Centre. ICNARC report on COVID-19 in critical care: England, Wales, and Northern Ireland [July 6, 2021]. Available at: https://www.icnarc.org/Our-Audit/Audits/Cmp/Reports. 2021.

12. Paediatric Intensive Care Audit Network report on COVID-19 confirmed cases in PICU (published 8th July 2021): Universities of Leeds and Leicester.

18. Bhalala US, Gist KM, Tripathi S, Boman K, Kumar VK, Retford L, et al. Characterization and Outcomes of Hospitalized Children With Coronavirus Disease 2019: A Report From a Multicenter, Viral Infection and Respiratory Illness Universal Study (Coronavirus Disease 2019) Registry. Crit Care Med. 2021.
19. Wearing HJ, Rohani P, Keeling MJ. Appropriate models for the management of infectious diseases. PLoS Med. 2005;2(7):e174.
20. Keeling MJ, Rohani P. Modeling Infectious Diseases in Humans and Animals: Princeton University Press; 2008.
21. Gaythorpe K, Imai N, Cuomo-Dannenburg G, Baguelin M, Bhatia S, Boonyasiri A, et al. Report 8: Symptom progression of COVID-19. Imperial College London COVID-19 Response Team. 2020.
22. Bi Q, Wu Y, Mei S, Ye C, Zou X, Zhang Z, et al. Epidemiology and transmission of COVID-19 in 391 cases and 1286 of their close contacts in Shenzhen, China: a retrospective cohort study. Lancet Infect Dis. 2020;20(8):911-9.
23. Herhalt C. Blood tests indicate one per cent of Ontario's population had COVID-19. CTV News. August 3, 2020. [Accessed September 11, 2020] Available at: https://toronto.ctvnews.ca/blood-tests-indicate-one-per-cent-of-ontario-s-population-had-covid-19-1.5049457.
24. Ontario Ministry of Health and Long-Term Care. COVID-19: Long-term care home surveillance testing and access to homes (Effective: July 16, 2021) Available at: https://www.ontario.ca/page/covid-19-long-term-care-home-surveillance-testing. July 16, 2021.
25. Kucirka LM, Lauer SA, Laeyendecker O, Boon D, Lessler J. Variation in False-Negative Rate of Reverse Transcriptase Polymerase Chain Reaction-Based SARS-CoV-2 Tests by Time Since Exposure. Ann Intern Med. 2020;173(4):262-7.
26. Carver C, Jones N. Comparative accuracy of oropharyngeal and nasopharyngeal swabs for diagnosis of COVID-19. Centre for Evidence-Based Medicine. 2020;Available at: https://www.cebm.net/covid-19/comparative-accuracy-of-oropharyngeal-and-nasopharyngeal-swabs-for-diagnosis-of-covid-19/. Epub March 26, 2020.
27. Ontario Ministry of Health and Long-Term Care. COVID-19 case data. [Accessed August 26, 2020] Available at: https://covid-19.ontario.ca/data.
28. Prem K, Cook AR, Jit M. Projecting social contact matrices in 152 countries using contact surveys and demographic data. PLoS Comput Biol. 2017;13(9):e1005697.
29. Public Health Ontario. COVID-19 Data and Surveillance. (Available at: https://www.publichealthontario.ca/en/data-and-analysis/infectious-disease/covid-19-data-surveillance).
30. Volz E, Mishra S, Chand M, Barrett JC, Johnson R, Geidelberg L, et al. Assessing transmissibility of SARS-CoV-2 lineage B.1.1.7 in England. Nature. 2021;593(7858):266-9.
31. Davies NG, Abbott S, Barnard RC, Jarvis CI, Kucharski AJ, Munday JD, et al. Estimated transmissibility and impact of SARS-CoV-2 lineage B.1.1.7 in England. Science. 2021;372(6538).
32. Yang W, Shaman J. Epidemiological characteristics of three SARS-CoV-2 variants of concern and implications for future COVID-19 pandemic outcomes. medRxiv. 2021:2021.05.19.21257476.
33. Scientific Pandemic Influenza Group on Modelling Operational sub-group (SPI-M-O). Consensus Statement on COVID-19 (June 2, 2021).
34. Zhang J, Litvinova M, Liang Y, Wang Y, Wang W, Zhao S, et al. Changes in contact patterns shape the dynamics of the COVID-19 outbreak in China. Science. 2020;368(6498):1481-6.
35. Webster RK, Brooks SK, Smith LE, Woodland L, Wessely S, Rubin GJ. How to improve adherence with quarantine: rapid review of the evidence. Public Health. 2020;182:163-9.
36. Stehle J, Voirin N, Barrat A, Cattuto C, Isella L, Pinton JF, et al. High-resolution measurements of face-to-face contact patterns in a primary school. PLoS One. 2011;6(8):e23176.
37. Salathe M, Kazandjieva M, Lee JW, Levis P, Feldman MW, Jones JH. A high-resolution human contact network for infectious disease transmission. Proc Natl Acad Sci U S A. 2010;107(51):22020-5.

38. Beutels P, Shkedy Z, Aerts M, Van Damme P. Social mixing patterns for transmission models of close contact infections: exploring self-evaluation and diary-based data collection through a web-based interface. Epidemiol Infect. 2006;134(6):1158-66.
39. Edmunds WJ, O'Callaghan CJ, Nokes DJ. Who mixes with whom? A method to determine the contact patterns of adults that may lead to the spread of airborne infections. Proc Biol Sci. 1997;264(1384):949-57.
40. Edmunds WJ, Kafatos G, Wallinga J, Mossong JR. Mixing patterns and the spread of close-contact infectious diseases. Emerg Themes Epidemiol. 2006;3:10.
41. Mikolajczyk RT, Kretzschmar M. Collecting social contact data in the context of disease transmission: prospective and retrospective study designs. Social Networks. 2008;30(2):127-35.
42. Najafi M, Laskowski M, de Boer PT, Williams E, Chit A, Moghadas SM. The Effect of Individual Movements and Interventions on the Spread of Influenza in Long-Term Care Facilities. Med Decis Making. 2017;37(8):871-81.
43. Champredon D, Najafi M, Laskowski M, Chit A, Moghadas SM. Individual movements and contact patterns in a Canadian long-term care facility. AIMS Public Health. 2018;5(2):111-21.
44. Roberts AR, Ishler KJ, Adams KB. The Predictors of and Motivations for Increased Family Involvement in Nursing Homes. The Gerontologist. 2018;60(3):535-47.
45. Angus Reid Institute. COVID-19 Compliance: One-in-five Canadians making little to no effort to stop coronavirus spread. August 17, 2020. Available at: http://angusreid.org/covid-compliance/.
46. Brooks JT, Butler JC. Effectiveness of Mask Wearing to Control Community Spread of SARS-CoV-2. JAMA. 2021;325(10):998-9. doi: 10.1001/jama.2021.1505. PubMed PMID: 33566056.
47. Chou R, Dana T, Jungbauer R. Update Alert 6: Masks for Prevention of Respiratory Virus Infections, Including SARS-CoV-2, in Health Care and Community Settings. Ann Intern Med. 2021. doi: 10.7326/L21-0393. PubMed PMID: 34251900; PubMed Central PMCID: PMCPMC8280537.
48. Mitze T, Kosfeld R, Rode J, Walde K. Face masks considerably reduce COVID-19 cases in Germany. Proc Natl Acad Sci U S A. 2020;117(51):32293-301.
49. Goolsbee A, Syverson C. Fear, Lockdown, and Diversion: Comparing Drivers of Pandemic Economic Decline 2020. National Bureau of Economic Research Working Paper Series. 2020;No. 27432. doi: 10.3386/w27432.
50. Care in Canadian ICUs. Canadian Institute for Health Information (CIHI). 2016.
51. Middlesex-London Health Unit. Summary of COVID-19 cases in Middlesex-London. Available at: https://www.healthunit.com/covid-19-cases-middlesex-london.
52. Province of Ontario. Ontario Makes COVID-19 Vaccination Policies Mandatory for High-Risk Settings. Press Release (August 17, 2021). Available at: https://news.ontario.ca/en/release/1000750/ontario-makes-covid-19-vaccination-policies-mandatory-for-high-risk-settings.
53. Pouwels KB, Pritchard E, Matthews PC, Stoesser N, Eyre DW, Vihta KD, et al. Impact of Delta on viral burden and vaccine effectiveness against new SARS-CoV-2 infections in the UK. Nuffield Department of Medicine, University of Oxford. 2021.
54. Pawlowski C, Lenehan P, Puranik A, Agarwal V, Venkatakrishnan AJ, Niesen MJM, et al. FDA-authorized mRNA COVID-19 vaccines are effective per real-world evidence synthesized across a multi-state health system. Med (N Y). 2021;2(8):979-92 e8.
55. Public Health England. COVID-19 vaccine surveillance report: 19 August 2021 (week 33) [Accessed August 20, 2021] Available at: https://www.gov.uk/government/publications/covid-19-vaccine-surveillance-report.
56. Tande AJ, Pollock BD, Shah ND, Farrugia G, Virk A, Swift M, et al. Impact of the COVID-19 Vaccine on Asymptomatic Infection Among Patients Undergoing Pre-Procedural COVID-19 Molecular Screening. Clin Infect Dis. 2021.
